# Durability of SARS-CoV-2 IgG Antibodies: Insights from a Longitudinal Study, Puerto Rico

**DOI:** 10.1101/2024.08.01.24311375

**Authors:** Zachary J. Madewell, Nathan Graff, Velma K. Lopez, Dania M. Rodriguez, Joshua M. Wong, Panagiotis Maniatis, Freddy A. Medina, Jorge L. Muñoz, Melissa Briggs-Hagen, Laura E. Adams, Vanessa Rivera-Amill, Gabriela Paz-Bailey, Chelsea G. Major

**Affiliations:** Division of Vector-Borne Diseases, Centers for Disease Control and Prevention, San Juan, Puerto Rico; Coronavirus and Other Respiratory Viruses Division, Centers for Disease Control and Prevention, Atlanta, Georgia; Division of Bacterial Diseases, Centers for Disease Control and Prevention, Atlanta, Georgia; Ponce Health Sciences University/Ponce Research Institute, Ponce, Puerto Rico

**Keywords:** Antibody dynamics, Omicron, Caribbean, COVID-19, vaccination, humoral immunity

## Abstract

Understanding the dynamics of antibody responses following vaccination and SARS-CoV-2 infection is important for informing effective vaccination strategies and other public health interventions. This study investigates SARS-CoV-2 antibody dynamics in a Puerto Rican cohort, analyzing how IgG levels vary by vaccination status and previous infection. We assess waning immunity and the distribution of hybrid immunity with the aim to inform public health strategies and vaccination programs in Puerto Rico and similar settings. We conducted a prospective, longitudinal cohort study to identify SARS-CoV-2 infections and related outcomes in Ponce, Puerto Rico, from June 2020–August 2022. Participants provided self-collected nasal swabs every week and serum every six months for RT-PCR and IgG testing, respectively. IgG reactivity against nucleocapsid (N) antigens, which generally indicate previous infection, and spike (S1) and receptor-binding domain (RBD) antigens, which indicate history of either infection or vaccination, was assessed using the Luminex Corporation xMAP® SARS-CoV-2 Multi-Antigen IgG Assay. Prior infection was defined by positive RT-PCRs, categorized by the predominant circulating SARS-CoV-2 variant at the event time. Demographic information, medical history, and COVID-19 vaccination history were collected through standardized questionnaires. Of 882 participants included in our analysis, 34.0% experienced at least one SARS-CoV-2 infection, with most (78.7%) occurring during the Omicron wave (December 2021 onwards). SARS-CoV-2 antibody prevalence increased over time, reaching 98.4% by the final serum collection, 67.0% attributable to vaccination alone, 1.6% from infection alone, and 31.4% from both. Regardless of prior infection status, RBD and S1 IgG levels gradually declined following two vaccine doses. A third dose boosted these antibody levels and showed a slower decline over time. N-antibody levels peaked during the Omicron surge and waned over time. Vaccination in individuals with prior SARS-CoV-2 infection elicited the highest and most durable antibody responses. N or S1 seropositivity was associated with lower odds of a subsequent positive PCR test during the Omicron period, with N antibodies showing a stronger association. By elucidating the differential decay of RBD and S1 antibodies following vaccination and the complexities of N-antibody response following infection, this study in a Puerto Rican cohort strengthens the foundation for developing targeted interventions and public health strategies.

## Introduction

The ongoing COVID-19 pandemic has spurred extensive research into diagnostic methods for current infections. Reverse transcription polymerase chain reaction (RT-PCR) and antigen tests are vital for detecting current infections, but retrospective identification of SARS-CoV-2 infections is also important for informing public health measures such as vaccine development and for understanding transmission dynamics. Serological assays offer rapid and reliable tools for identifying prior infections by detecting antibodies following SARS-CoV-2 infection.^1,2^ These assays commonly target immunoglobulins M (IgM) and G (IgG) antibodies in humans.^3^

IgG antibodies are generally detectable for longer periods after infection compared to IgM and potentially play a role in long-term immunity following SARS-CoV-2 infection or vaccination. During the human immune response to SARS-CoV-2, IgG conversion typically occurs around 14 days post-infection, with antibodies remaining detectable for up to at least 15 months.^4–7^ Recent studies have revealed variations in immune responses to COVID-19 vaccines, with mRNA vaccines eliciting higher antibody affinity and resulting IgG titers compared to other vaccine types.^8–10^ Receiving three or more doses of mRNA vaccines have been shown to provide greater IgG durability than completion of just two vaccine doses,^9^ offering strong protection against hospitalization with vaccine effectiveness estimates of 82.5% (95% CI: 77.8%–86.2%) after the third vaccine dose and 87.3% (95% CI: 75.5%–93.4%) after the fourth dose.^11^ Hybrid immunity, resulting from both previous infection and vaccination, has been reported to provide better protection compared to infection or vaccination alone.^12^ Serological assays that detect IgG antibodies against multiple SARS-CoV-2 proteins can be valuable in differentiating between previous infection and vaccination, as well as assessing potential differences in resulting immunity duration. However, knowledge gaps remain regarding the longevity of these protective responses, particularly with the emergence of new variants.

Key IgG targets include the nucleoprotein (N) and the spike (S1) glycoprotein’s receptor-binding domain (RBD), both major structural viral proteins of SARS-CoV-2.^13–15^ S1 interacts with the human ACE2 receptor, facilitating viral entry, whereas RBD is a specific binding site within S1 targeted by neutralizing antibodies.^16^ Understanding the persistence of these antibodies post-infection is essential for evaluating immunity, especially in the context of vaccination. S1 and RBD antibodies may indicate both infection-induced and vaccine-induced immunity. Conversely, N antibodies primarily indicate infection-induced immunity due to the exclusion of N protein in current FDA-approved vaccines which focus on eliciting an immune response that targets S1. In settings with high vaccination rates, N antibody assays can be helpful in distinguishing prior SARS-CoV-2 infections from vaccination-induced responses, especially for mild or asymptomatic infections which often go undetected during their acute phase.^17^

Waning of infection-induced or vaccine-induced antibodies over time is a concern, particularly with the emergence of new SARS-CoV-2 variants.^18,19^ Different variants may elicit diverse antibody responses and be variably affected by immunity from prior infections or vaccination. For example, infection with the Omicron variant has been shown to result in a higher anti-N IgG response than other variants, particularly among vaccinated individuals.^20^ The Omicron variant showed greater resistance to IgG neutralization generated from early (wild-type) infection or original COVID-19 monovalent vaccines, leading to relatively high infection rates in previously infected and vaccinated individuals.^6,21^ In light of these findings, further characterization of immune response durability is necessary. This includes investigating how factors such as age, underlying health conditions, and the combined effect of vaccination and infection history influence IgG persistence and resulting immunity.

This study investigates SARS-CoV-2 IgG antibody responses (N, S1, RBD) in a community-based cohort in Puerto Rico during 2020–2022. We analyze IgG levels by vaccination status and previous infection, assessing potential waning over time and variations related to age, chronic conditions, putative SARS-CoV-2 variant based on collection time, and vaccination-infection sequence. Additionally, we estimate the distribution of hybrid immunity over time and evaluate associations between IgG seroreactivity and subsequent infections. These analyses aim to provide insights into long-term immunity against SARS-CoV-2, informing public health strategies and vaccination programs in Puerto Rico and similar settings.

## Methods

### Study Design and Population

Communities Organized to Prevent Arboviruses (COPA) is an ongoing cohort study launched in 2018 in Ponce, Puerto Rico, to assess arbovirus burden and control interventions in a community-based population. COPA is a collaboration involving Ponce Health Sciences University/Ponce Research Institute, the Puerto Rico Vector Control Unit, and the U.S. Centers for Disease Control and Prevention (CDC). Study enrollment and data collection activities are described elsewhere.^22,23^ Approval for the COPA project was obtained from the Ponce Medical School Foundation, Inc. Institutional Review Board (protocol number 2402185168/171110-VR).

A COPA sub-study for COVID-19, COCOVID, was implemented in June 2020, enrolling COPA participants and other residents from 15 selected community areas.^24,25^ Eligible individuals were ≥1 year old, spent four or more nights a week in the selected residence, and had no definite plans to move in the next 12 months. Primary study enrollment occurred from June 2020 to February 2021, with subsequent secondary enrollment until April 2022 limited to household members of active participants. Participants answered questionnaires and provided serum for multi-IgG antibody testing at enrollment and every 6 months until August 2022. They also provided weekly self-collected anterior nasal swabs for SARS-CoV-2 RT-PCR testing until April 2022. Additional anterior nasal swabs for expedited RT-PCR testing were collected by study staff for participants with COVID-like symptoms or close contact with a COVID-19 case during the entire COCOVID study period and until February 2023 for all participants that remained active in the COPA cohort.

### IgG Antibody Testing

The Luminex xMAP® SARS-CoV-2 Multi-Antigen IgG Assay^26^ was used according to the manufacturer’s instructions to assess seroreactivity in COCOVID serum specimens against three key antigen components: N, S1, and RBD.^27^ The multiplex assay uses Luminex MagPlex beads for each IgG antigen target, and specimens were considered positive for N, S1, or RBD when their median fluorescence intensity (MFI) was >700 call threshold. The MFI is considered a measure of the total degree of saturation in the Luminex test platform and was used as a surrogate marker for the level of antibody titers in our analyses.

### Infection Events and Status

SARS-CoV-2 infection was defined by positive RT-PCR results from nasal swabs collected weekly throughout the study period (June 2020–August 2022), including up to 6 months after the participants’ final serological test. In our analyses, participant infection status at each serum collection date was assigned based on RT-PCR test results from all previously collected swabs, either as having had one or more previous RT-PCR positive results or all negative previous RT-PCR results. Positive tests separated by ≥90 days and accompanied by at least one negative test in between were considered separate illness episodes.^28^

Based on dominant variants circulating in Puerto Rico, infection events that occurred prior to and after serum was collected for IgG testing were categorized by collection date of first positive nasal swab: pre-Delta (June 2020–May 2021), Delta (June–November 2021), and Omicron (December 2021–February 2023)^29,30^; pre-Delta variant circulation periods were not individually defined due to low sample size. For participants with more than one apparent infection event within a 6-month period, the variant circulation period for the most recent infection was assigned.

### Vaccination Status, Hybrid Immunity, and Other Participant Characteristics

Questionnaires answered by participants at enrollment and every 6 months during follow-up included data collection on demographics, health conditions, and COVID-19 vaccination status, including the number of doses, date of each dose, and manufacturer (Pfizer-BioNTech’s BNT162b2, Moderna’s mRNA-1273). Serum specimens were categorized based on the number of COVID-19 vaccine doses the participant had received prior to the collection date: unvaccinated (no doses), 1 dose, 2 doses, and 3 doses.

Consistent with previous research,^31^ for the sub-analysis of hybrid immunity, vaccine-induced immunity was defined as history of at least one COVID-19 vaccination confirmed by vaccination cards, infection-induced immunity was defined as anti-N and anti-S1 antibody detection, and hybrid immunity was defined as combined protection from ≥1 dose of COVID-19 vaccination, and anti-N and anti-S1 antibody detection.

Sera were also categorized by participant age group at the time of collection (<20 years, 20–39 years, 40–64 years, ≥65 years), and whether they had been previously diagnosed with any chronic health condition by a medical provider. Any chronic health condition included reporting one or more previously diagnosed physical or mental health conditions, and serum specimens from participants with and without specific physical conditions, including hypertension, chronic respiratory disease, diabetes, and high triglycerides, were also compared.

### Sample Population

We analyzed blood samples (serum specimens) collected at 6-, 12-, and 18-month follow-up visits from participants in the COCOVID study (December 2020–August 2022). Blood samples from enrollment and the 24-month visit were excluded due to incomplete data on past infections using RT-PCR tests. Weekly anterior nasal swab collections for all participants ended in April 2022, before any participant reached their 24-month visit. However, RT-PCR results from anterior nasal swabs up to the 18-month visit were used to categorize IgG responses. All RT-PCR results throughout the study period, including weekly anterior nasal swabs up to April 2022 and additional swabs collected for symptomatic testing or close contact investigation, were considered to identify infections that occurred after vaccination or a documented prior infection. Blood samples from 6-, 12-, and 18-month visits were included if participants met one of the following criteria: 1) completed at least 80% of RT-PCR tests within the preceding 6 months, or 2) had a positive RT-PCR test result within the preceding 6 months. Due to the low number of participants receiving Johnson & Johnson’s single-dose Ad26.COV2.S vaccine in this cohort, analyses excluded serum specimens from those who only received this vaccine. However, participants who received at least one dose of an mRNA vaccine after an initial Ad26.COV2.S dose were included.

### Statistical Analysis

The prevalence of vaccine-induced, infection-induced, or hybrid immunity was estimated for four time periods (Dec 2020–Apr 2021, May–Sep 2021, Oct 2021–Feb 2022, Mar–Aug 2022), stratified by age group. SARS-CoV-2 IgG anti-N, anti-RBD, and anti-S1 responses were assessed by days since last COVID-19 vaccine dose and vaccination status, days since RT-PCR positivity, and sequence of RT-PCR positivity and vaccination for those completing a two-dose primary series. Medians and interquartile ranges (IQR) for the MFI were calculated for each of the three IgG responses by vaccination status, days since vaccination (<14, 14–27, 28–89, 90–179, ≥180), and months since RT-PCR positivity (<2, 2–4, ≥5, no previous positive RT-PCR). All MFI values were log-transformed. For participants without prior RT-PCR positivity and negative anti-N antibodies, we evaluated anti-S1 and anti-RBD seropositivity by days since their last vaccine. Anti-N seropositivity rates were examined in previously infected individuals by vaccination status.

Generalized linear mixed-effects regression, with participant ID included as a random effect, was used to evaluate unadjusted and adjusted associations between anti-N, anti-RBD, and anti-S1 IgG responses and time since RT-PCR positivity, time since last vaccine dose, and chronic conditions (see Appendix S1 for additional details). Logistic regression with a random intercept was used to analyze associations between IgG seroreactivity and RT-PCR confirmed infection in subsequent six months during the Omicron predominant period. All available data from participants at each follow-up visit was included in the analysis, regardless of their subsequent participation status. All analyses were done using R software, version 4.3.1 (R Foundation for Statistical Computing, Vienna, Austria).

## Results

### Study Population

Among the 1,030 participants enrolled in the COCOVID study, 882 (85.6%) had one or more serologic test results included in our analyses. Of these 882 participants, 789 (89.5%) had qualifying serologic test results available at 6 months, 774 (87.8%) at 12 months, and 567 (64.3%) at 18 months following study enrollment (Table S1). Median age of these participants at baseline was 37 years (IQR: 18–49), 52.7% were female, and 59.3% had any chronic health condition, of which hypertension (22.4%), respiratory illness (17.2%), and diabetes (11.6%) were most frequently reported (Table 1). By their final serologic test, 86 (9.8%) participants were unvaccinated, 10 (1.1%) had received one dose, 316 (35.8%) had received two vaccine doses, 468 (53.1%) had received three doses, and two (0.2%) had received four doses (Tables 1, S1).

**Table 1.** Descriptive characteristics of COCOVID participants, Puerto Rico, 2020–2022.

| Characteristic | N = 882<br>n (%) |
| --- | --- |
| **Age group** |  |
| <20 years | 249 (28.2) |
| 20–39 years | 237 (26.9) |
| 40–64 years | 300 (34.0) |
| ≥65 years | 96 (10.9) |
| **Sex** |  |
| Female | 465 (52.7) |
| Male | 417 (47.3) |
| **Any chronic condition<sup>a</sup>** |  |
| Yes | 523 (59.3) |
| No | 359 (40.7) |
| **Follow-up visit** |  |
| 6 months | 789 (37.0) |
| 12 months | 774 (36.3) |
| 18 months | 567 (26.6) |
| **COVID-19 vaccination status recorded on final visit** |  |
| Unvaccinated | 86 (9.8) |
| One dose | 10 (1.1) |
| Two doses | 316 (35.8) |
| Three doses | 468 (53.1) |
| Four doses | 2 (0.2) |
| **Positive SARS-CoV-2 RT-PCR during study** |  |
| Any positive RT-PCR | 300 (34.0) |
| Pre-Delta predominant period | 28 (3.2) |
| Delta predominant period | 17 (1.9) |
| Omicron predominant period | 263 (29.8) |
| No positive RT-PCR | 582 (66.0) |

<sup>a</sup> Any chronic condition included hypertension (N=198), chronic respiratory illness (N=152), diabetes (N=102), cardiac conditions (N=80), thyroid conditions (N=81), gastrointestinal problems (N=75), high triglycerides (N=72), asthma (N=60), arthritis (N=57), neurological disorders (N=35), anxiety (N=33), rheumatic conditions (N=28), depression (N=27), blood disorder (N=24), fibromyalgia (N=24), kidney conditions (N=24), cancer (N=23), mental illness (N=17), physical disabilities (N=15), liver conditions (N=14), obesity (N=14), stroke (N=9), and other (N=155).

### SARS-CoV-2 Infection

Of the 882 participants with one or more serologic test results, 300 (34.0%) had at least one positive SARS-CoV-2 RT-PCR test during the study period, with eight (2.7%) experiencing two separate RT-PCR confirmed infections. By the 18-month serologic test, 20.7% (183/882) had at least one positive RT-PCR test, with 78.7% (n=144) occurring during the Omicron predominant period (December 2021 onwards). The remaining 119 infections occurred in the 6 months following the final serologic test, all of which occurred during the Omicron predominant period.

### Distribution of Vaccine-induced, Infection-induced, and Hybrid Immunity over Time

The prevalence of SARS-CoV-2 antibodies indicative of prior infection or vaccination (both anti-N and anti-S1 positive or anti-N positive only) increased substantially over the study period. In the earliest analysis period (December 2020–April 2021), only 26.9% (95% CI: 23.2%, 30.9%) of 528 participants with serological test results had detectable SARS-CoV-2 antibodies. This included individuals with vaccine-induced immunity alone (25.2%), infection-induced immunity alone (0.9%), and hybrid immunity (0.8%) (Figure 1). By the final period (March–August 2022), 98.4% (95% CI: 96.6%, 99.3%) of 564 participants with eligible results had SARS-CoV-2 antibodies, with 67.0% from vaccination alone, 1.6% from infection alone, and 31.4% from both. During this final period, the prevalence of hybrid immunity was highest among participants <20 years (39.3%, 95% CI: 31.1%, 48.1%) and lowest among those ≥65 years (13.0%, 95% CI: 5.4%, 27.0%).

**Figure 1.**
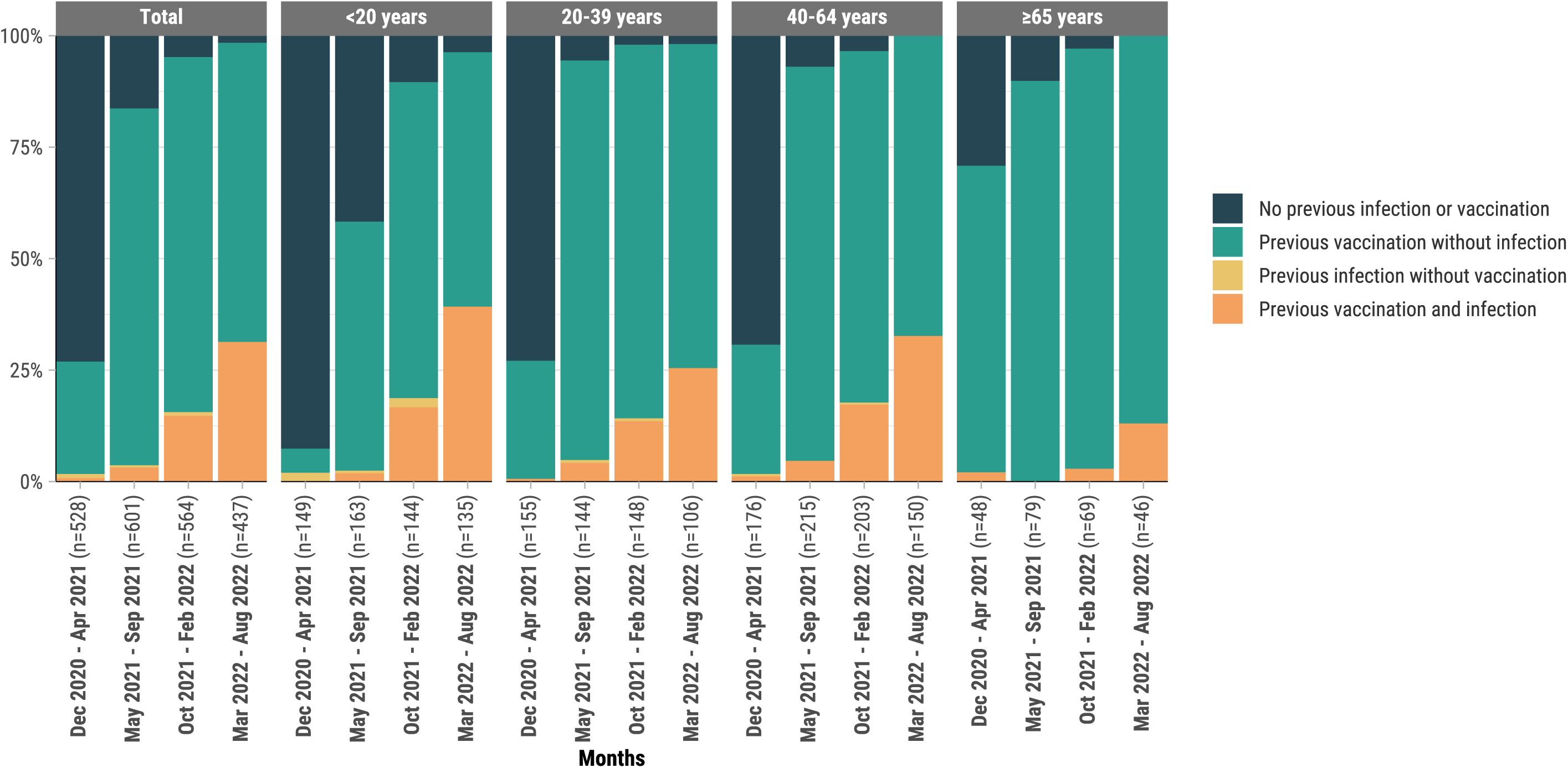
Distribution of vaccine-induced, infection-induced, and hybrid (immunity derived from a combination of vaccination and infection) immunity^a^ against SARS-CoV-2 by age group, Puerto Rico, 2020–2022. ^a^Combined detection of anti-S1 antibodies (produced by both COVID-19–vaccination and SARS-CoV-2 infection) and anti-N antibodies (specific to prior infection), along with vaccination history from vaccination cards. *Vaccine-induced immunity*: Individuals with history of ≥1 COVID-19 vaccination dose from vaccination card. *Infection-induced immunity*: Individuals with positive anti-N antibodies and positive anti-S1 antibodies. *Hybrid immunity*: Individuals with self-reported history of ≥1 COVID-19 vaccination dose and positive antibodies for both S1 and N antigens.

The widespread availability of COVID-19 vaccines in Puerto Rico (April–May 2021) coincided with an increase in RBD and S1 antibody titers (Figure 2). Among participants previously negative for RT-PCR and N antibodies, all who had a serum sample collected within 7–13 days post-second vaccine dose seroconverted for both S1 and RBD antibodies. However, by 90–179 days after the second dose, S1 detection dropped to 75.6%, while RBD remained detectable in 99.1% (Figures S1, S2). A third vaccine dose resulted in 100% detection of both RBD and S1 antibodies at 90–179 days post-vaccination.

**Figure 2.**
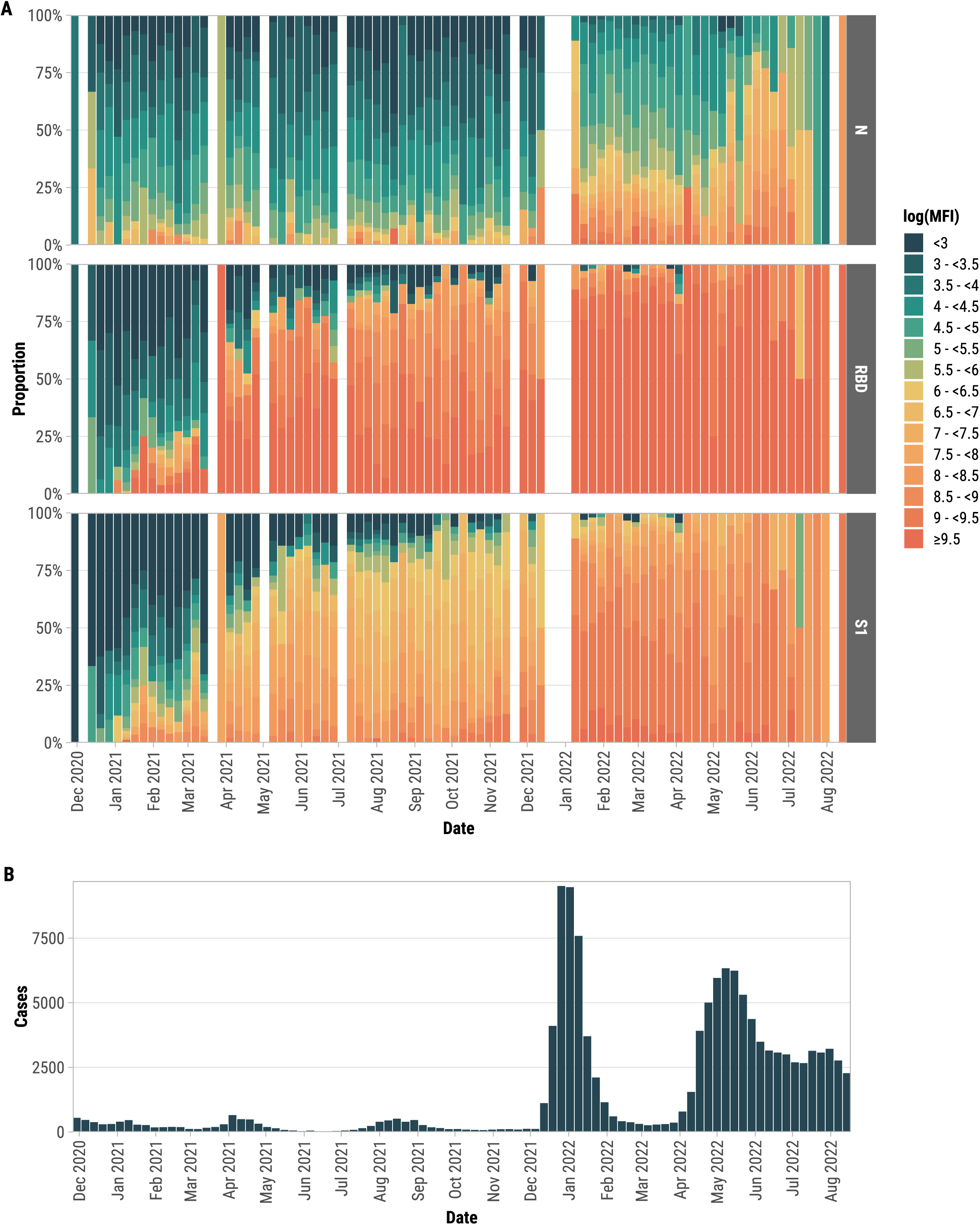
(A) Weekly distribution of SARS-CoV-2 N, RBD, and S1 antibody responses during the serosurvey period and B) case counts (confirmed and probable) reported to the Puerto Rico Department of Public Health in Ponce Health Region, 2020–2022. Departamento de Salud PR. COVID-19 en cifras en Puerto Rico. 2024 [cited 2024 May 17]; Available from: https://www.salud.pr.gov/estadisticas_v2.

N antibody levels increased dramatically during a surge of COVID-19 cases in Puerto Rico following the introduction of the Omicron variant at the end of 2021 (Figure 2). Among participants tested during this period, 28.3% (196/692) showed N IgG seroconversion, of whom 75.5% (148/196) had a previous positive RT-PCR test. Among participants who seroconverted for N antibodies, median log(MFI) for N antibodies was higher in participants with a previous positive RT-PCR (8.20, IQR: 7.52, 8.70) than those with negative RT-PCRs (7.60, IQR: 7.24, 8.61) regardless of vaccination status (*p*=0.022).

### IgG Responses and Waning by Vaccination Status

Analysis of all serum samples suggests that RBD and S1 antibody responses were more commonly detected and persisted longer than N antibody responses across all vaccination groups (Figures 3, S3). For individuals who received two doses, the overall median log(MFI) at any time post-vaccination was 9.42 (IQR: 8.93, 9.70) for RBD, 7.55 (IQR: 6.86, 8.17) for S1, and 4.05 (IQR: 3.37, 4.87) for N (Figure S3). RBD and S1 antibodies were highly correlated regardless of vaccine dosage (*r* ≥ 0.83) (Figure S4).

**Figure 3.**
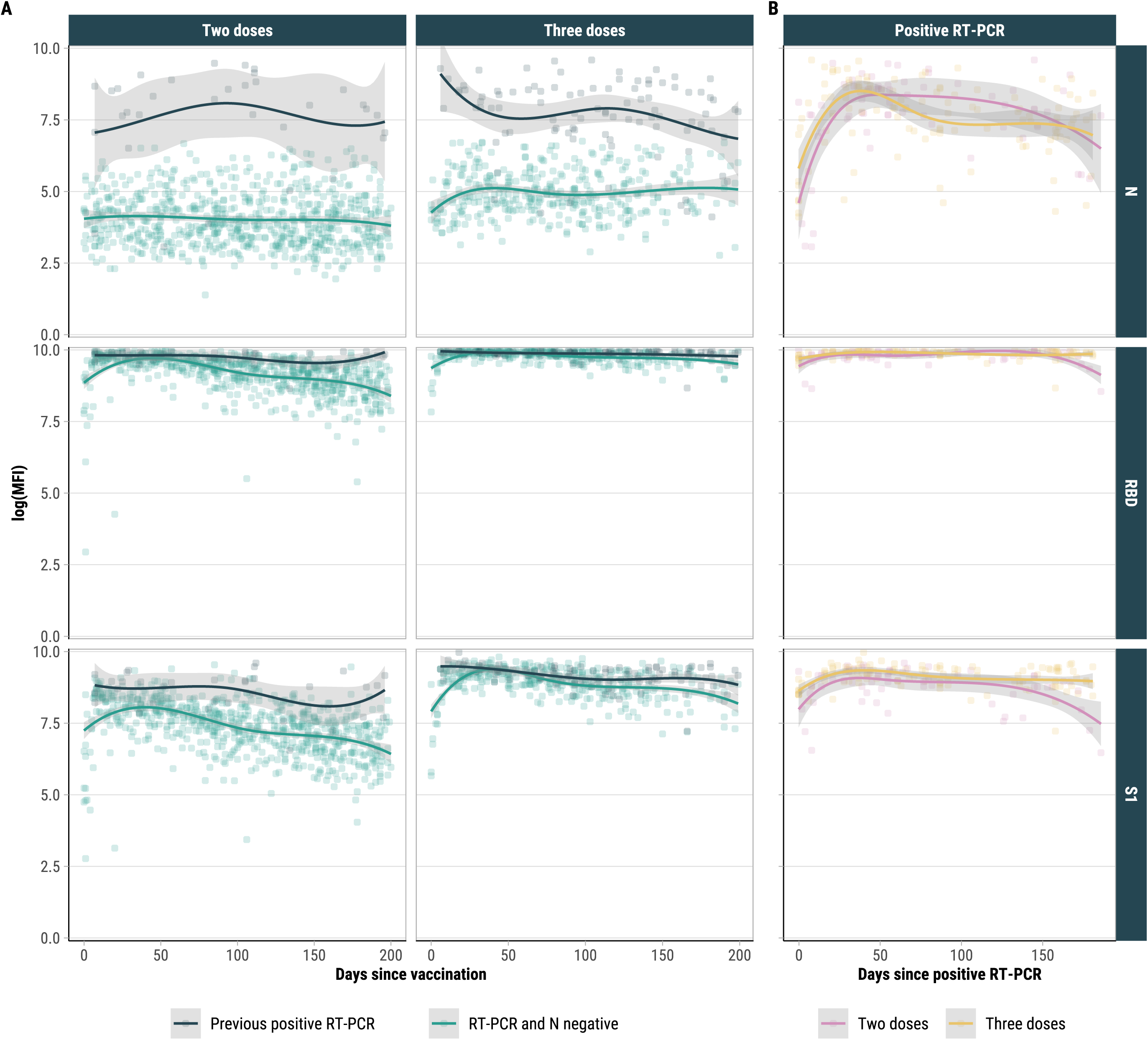
SARS-CoV-2 IgG antibody responses by days since last vaccine stratified by RT-PCR positivity in previous six months vs. no previous RT-PCR positivity and N negative (A) and antibody responses by days since last positive RT-PCR and vaccination status (B), Puerto Rico, 2020–2022. The lines are loess smoothing lines and shaded bands are 95% CIs.

In participants who received two vaccine doses without a prior positive RT-PCR, both RBD and S1 levels gradually waned over time (Figures 3, S3). The median log(MFI) for RBD and S1 decreased from 9.73 (IQR: 9.64, 9.83) and 8.09 (IQR: 7.73, 8.38) at 14–27 days post-vaccination to 8.65 (IQR: 8.24, 9.12) and 6.64 (IQR: 6.16, 7.26) at ≥180 days, respectively. Younger participants (<20 years) had higher antibody levels than those aged ≥65 at later time points (>180 days post-vaccination), with a median log(MFI) for S1 of 7.51 versus 6.35, and for RBD, 9.22 versus 8.48 (Figure S5).

Almost all antibody tests for participants who received a third vaccine dose were conducted during the Omicron predominant period. Among all serum samples collected from participants with a third dose, both RBD and S1 antibody levels peaked at 14–27 days post-vaccination (median log[MFI]: 9.90, IQR: 9.85, 9.94, and 9.22, IQR: 9.05, 9.47, respectively), declining thereafter to 9.74 (IQR: 9.48, 9.90) and 8.79 (IQR: 8.33, 9.08) ≥180 days post-vaccination, respectively (Figure S3). Adjusting for age group, sex, chronic conditions, and previous infection, log(MFI) for RBD and S1 decreased by -0.002 (95% CI: -0.004, -0.001) and -0.009 (95% CI: -0.012, -0.005), respectively, for each month post-third dose (Figure S6). RBD and S1 responses were higher for participants who received a third dose than those who with second doses regardless of age group or time since vaccination (Figure S5).

To directly assess the rate of antibody decline, we analyzed changes in IgG levels between paired serum samples collected at different time points after the second and third vaccine doses in participants with and without prior PCR-confirmed infection. For individuals with a second dose and without a previous positive RT-PCR, RBD and S1 levels fell by 8.8% (from 9.73 to 8.78) and 20.9% (from 8.44 to 6.68), respectively, after six months (Figure S7). For those with a third dose and without a previous positive RT-PCR, RBD and S1 levels fell by 2.3% (from 9.82 to 9.59) and 6.2% (from 8.97 to 8.41), respectively, after six months (Figure 4). For those with a third dose and a previous positive RT-PCR, S1 levels decreased by 4.1% (from 9.21 to 8.83), while RBD levels remained stable at 9.90 after six months. There were no participants with paired SARS-CoV-2 IgG antibody responses at two time points six months apart with a second dose and a previous positive RT-PCR. N-specific antibodies for sample pairs demonstrate N levels fell by 6.9% (from 7.84 to 7.30) for those with a positive RT-PCR.

**Figure 4.**
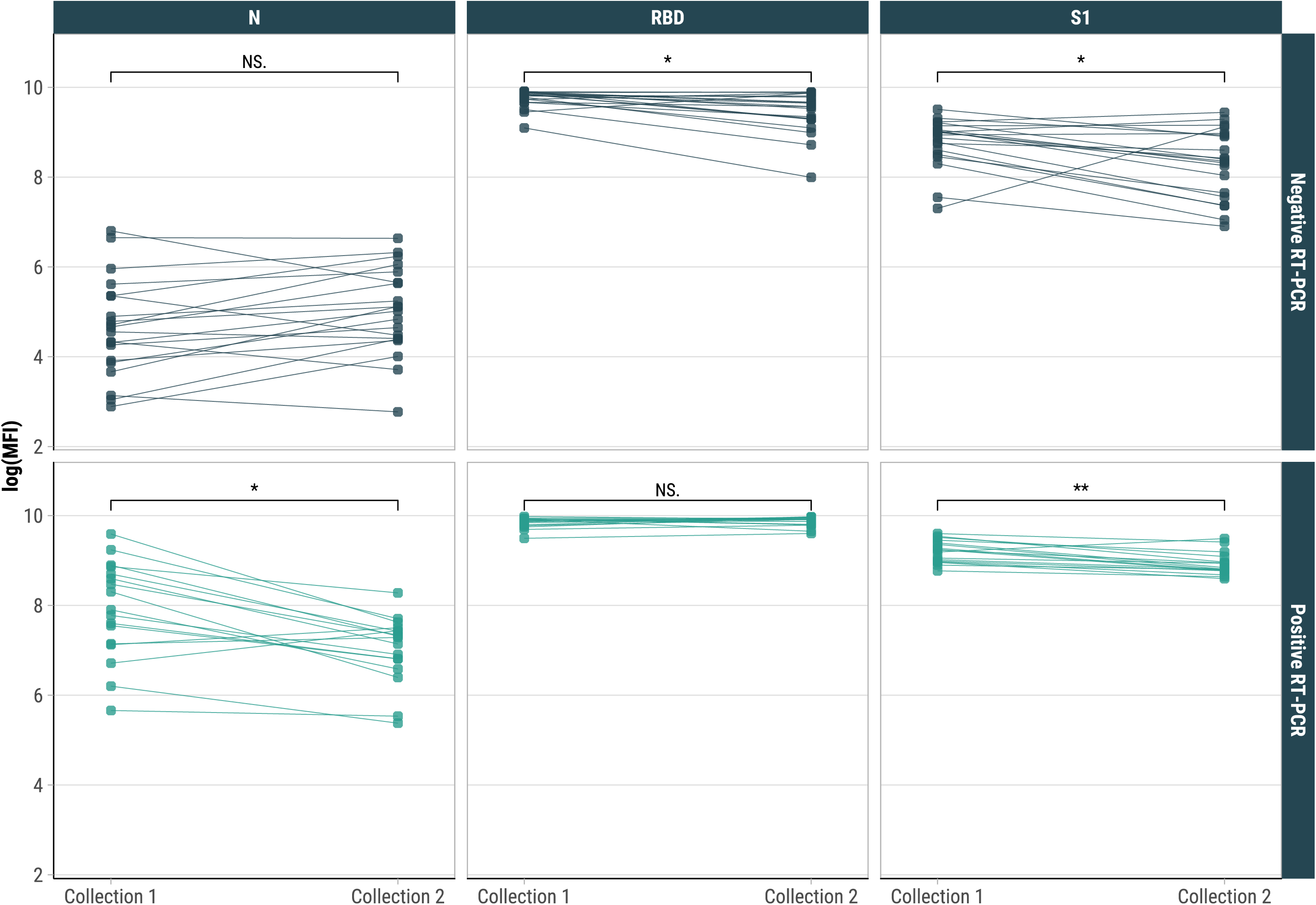
Paired SARS-CoV-2 IgG antibody responses for individuals at two time points six months apart among people who had received three vaccine doses by prior RT-PCR positivity, Puerto Rico, 2020–2022. \**P*_<_0.05, \*\**P* < 0.01. NS: not significant.

### Nucleocapsid Responses and RT-PCR Positivity

Among individuals with a previous positive RT-PCR (80.8% N seropositive), N antibody levels were higher (median log[MFI]: 7.89, IQR: 6.90, 8.56) compared to those without a positive RT-PCR (median log[MFI]: 4.34, IQR: 3.60, 5.13) (Figures 3, S3). N antibody levels peaked within two months of a positive test (median log[MFI]: 8.27, IQR: 7.09, 8.69) and gradually declined thereafter, reaching a median log(MFI) of 7.24 (IQR: 6.43, 8.30) at five months after testing positive. Among individuals without a history of previous infection, N-seropositivity following an infection was 83.7% and 74.6% for those who received two and three doses, respectively, not significantly different from unvaccinated individuals (77.8%) (*p*=0.498) (Table 2).

**Table 2.** Anti-N seropositivity rates among participants who tested positive for SARS-CoV-2 by RT-PCR at least 14 days after vaccination and were previously seronegative and RT-PCR negative, Puerto Rico, 2020–2022.

| Vaccination status | All<br>Anti-N |  | Pre-Omicron<br>Anti-N |  | Omicron<br>Anti-N |  |
| --- | --- | --- | --- | --- | --- | --- |
|  | Participants<br>N | seropositivity<br>% (95% CI) | Participants<br>N | seropositivity<br>% (95% CI) | Participants<br>N | seropositivity<br>% (95% CI) |
| Unvaccinated | 18 | 77.8 (51.9, 92.6) | 16 | 81.2 (53.7, 95.0) | 2 | 50.0 (9.5, 90.5) |
| Two doses | 49 | 83.7 (69.8, 92.2) | 13 | 69.2 (38.9, 89.6) | 36 | 88.9 (73.0, 96.4) |
| Three doses | 67 | 74.6 (62.3, 84.1) | – | – | 67 | 74.6 (62.3, 84.1) |

Both vaccination and previous RT-PCR positivity provided a stronger boost to RBD and S1 antibody responses compared to either factor alone (Figures 3, S3). The order of vaccination and RT-PCR positivity did not significantly impact antibody responses among participants who received two doses (Figure S8).

### IgG Seroreactivity and Subsequent RT-PCR positivity

Our analysis revealed that individuals with detectable antibodies against either N or S1 were less likely to experience a positive RT-PCR test in the following six months during the Omicron-dominant period. Specifically, N and S1 seropositivity were associated with 90.3% (95% CI: 78.8%, 95.6%) and 58.3% (95% CI: 12.1%, 80.2%) lower odds, respectively, of a subsequent positive test, after adjusting for age group, sex, and chronic conditions (Table 3). Participants with a higher N antibody level (log(MFI) ≥ 8.55) had an even greater reduction in odds (96.3%, 95% CI: 73.0%, 99.5%) compared to those who were N seronegative. These findings suggest that SARS-CoV-2 infection, as indicated by N seropositivity, provided substantial protection against reinfection during the Omicron period, while vaccination, as indicated by S1 seropositivity, also offered significant, though comparatively lower, protection.

**Table 3.**
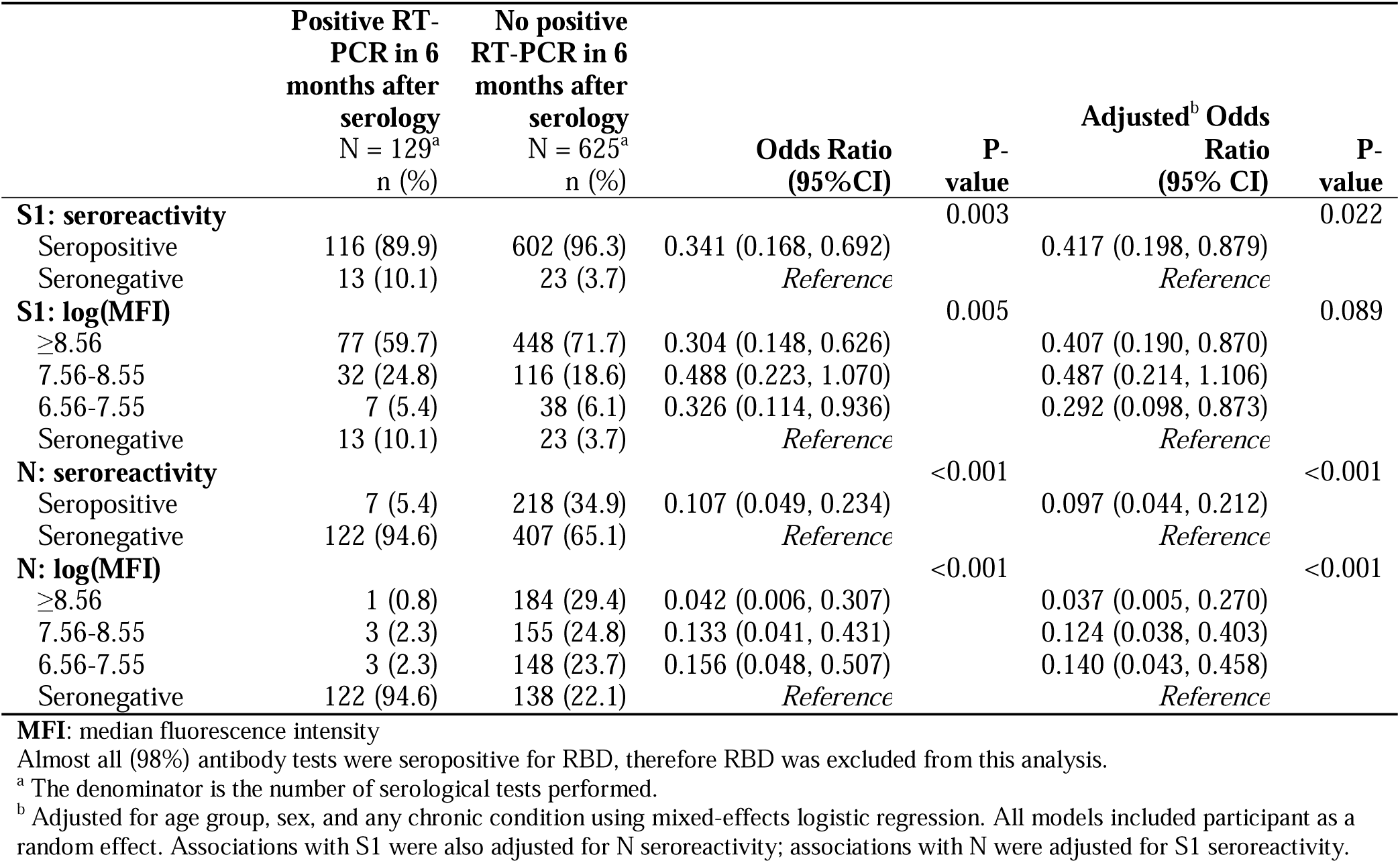
Associations between SARS-CoV-2 seroreactivity and subsequent RT-PCR positivity within six months during the Omicron predominant period, Puerto Rico, 2020–2022.

|  | Positive RT-PCR in 6 months after serology<br>N = 129<sup>a</sup><br>n (%) | No positive RT-PCR in 6 months after serology<br>N = 625<sup>a</sup><br>n (%) | Odds Ratio<br>(95% CI) | P-value | Adjusted<sup>b</sup> Odds Ratio<br>(95% CI) | P-value |
| --- | --- | --- | --- | --- | --- | --- |
| **S1: seroreactivity** |  |  |  | 0.003 |  | 0.022 |
| Seropositive | 116 (89.9) | 602 (96.3) | 0.341 (0.168, 0.692) |  | 0.417 (0.198, 0.879) |  |
| Seronegative | 13 (10.1) | 23 (3.7) | *Reference* |  | *Reference* |  |
| **S1: log(MFI)** |  |  |  | 0.005 |  | 0.089 |
| ≥8.56 | 77 (59.7) | 448 (71.7) | 0.304 (0.148, 0.626) |  | 0.407 (0.190, 0.870) |  |
| 7.56-8.55 | 32 (24.8) | 116 (18.6) | 0.488 (0.223, 1.070) |  | 0.487 (0.214, 1.106) |  |
| 6.56-7.55 | 7 (5.4) | 38 (6.1) | 0.326 (0.114, 0.936) |  | 0.292 (0.098, 0.873) |  |
| Seronegative | 13 (10.1) | 23 (3.7) | *Reference* |  | *Reference* |  |
| **N: seroreactivity** |  |  |  | <0.001 |  | <0.001 |
| Seropositive | 7 (5.4) | 218 (34.9) | 0.107 (0.049, 0.234) |  | 0.097 (0.044, 0.212) |  |
| Seronegative | 122 (94.6) | 407 (65.1) | *Reference* |  | *Reference* |  |
| **N: log(MFI)** |  |  |  | <0.001 |  | <0.001 |
| ≥8.56 | 1 (0.8) | 184 (29.4) | 0.042 (0.006, 0.307) |  | 0.037 (0.005, 0.270) |  |
| 7.56-8.55 | 3 (2.3) | 155 (24.8) | 0.133 (0.041, 0.431) |  | 0.124 (0.038, 0.403) |  |
| 6.56-7.55 | 3 (2.3) | 148 (23.7) | 0.156 (0.048, 0.507) |  | 0.140 (0.043, 0.458) |  |
| Seronegative | 122 (94.6) | 138 (22.1) | *Reference* |  | *Reference* |  |

**MFI:** median fluorescence intensity
Almost all (98%) antibody tests were seropositive for RBD, therefore RBD was excluded from this analysis.
<sup>a</sup> The denominator is the number of serological tests performed.
<sup>b</sup> Adjusted for age group, sex, and any chronic condition using mixed-effects logistic regression. All models included participant as a random effect. Associations with S1 were also adjusted for N seroreactivity; associations with N were adjusted for S1 seroreactivity.

### Chronic Conditions and IgG Responses

Among participants who received two doses, diabetes was associated with lower RBD (β: -0.022, 95% CI: -0.039, -0.006) and S1 (β: -0.029, 95% CI: -0.056, -0.002) responses compared to those without diabetes, after adjusting for age group, sex, and previous RT-PCR positivity (Figure S9). Similarly, for participants who received a third dose, high triglycerides were associated with lower RBD (β: -0.009, 95% CI: -0.016, -0.003) and S1 (β: -0.024, 95% CI: -0.040, -0.007) responses compared to those with normal triglyceride levels. No significant associations were found between other chronic conditions and antibody responses.

## Discussion

This study provides novel real-world data on SARS-CoV-2 antibody dynamics in a semi-isolated Caribbean island population throughout the COVID-19 pandemic. This unique setting allows for a more controlled analysis of antibody response compared to geographically dispersed populations. We observed a substantial rise in seropositivity (from 26.9% to 98.4%) over the study period, including an increase in hybrid immunity from 0.8% to 31.4%. Studies have shown that hybrid immunity offers several advantages, including enhanced protection against severe disease and death, slower waning of immunity, reduced risk of reinfection, promotion of B cell diversity, and facilitation of T cell activation during subsequent vaccination.^32^ However, the impact of this increase on population immunity remains unclear due to the emergence and potential immune escape of SARS-CoV-2 variants. These findings emphasize the importance of ongoing surveillance and tailored vaccination strategies to effectively manage the COVID-19 pandemic.

RBD and S1 antibody levels gradually declined over time following initial vaccine doses, reflecting the natural decay of vaccine-induced humoral immunity. S1 levels waned faster than RBD levels, consistent with previous research.^33^ This difference might be due to the inherent structural stability of the RBD region,^34^ potentially contributing to slower degradation and potentially longer-lasting protection against viral entry. However, limitations of the assay used warrant further investigation with assays optimized for S1 and RBD detection to definitively assess their relative decay rates. As expected, booster doses substantially increased RBD and S1 levels compared to the primary series, peaking 14-27 days post-booster dose, indicating a rapid and robust immune response.

Recent SARS-CoV-2 infections boosted N, RBD, and S1 antibody levels, peaking within two months and gradually declining after six months, suggesting natural humoral immunity decay. Although infection may provide protective benefits through antibody boosts, vaccination remains the safest strategy for building immunity against COVID-19, considering the risk of severe illness during infection.

One-quarter of participants with N seroconversion during the Omicron-predominant period lacked a positive RT-PCR result. This could be due to asymptomatic or mild infections, transient viral RNA levels below the detection threshold, or infections that were too recent for RT-PCR detection. Alternatively, they could have had an acute infection that was not yet RT-PCR detectable. Supporting this hypothesis, a previous study reported declining RT-PCR sensitivity for detecting asymptomatic cases over time, with peak sensitivity occurring within the first few days after infection.^35^ Conversely, some participants with positive RT-PCRs had negative N tests, possibly reflecting early infection before significant antibody development. Alternatively, some individuals with mild or asymptomatic infections might not generate a strong immune response. As a result, their antibody production, including N antibodies, may be minimal or undetectable.^36^ These findings highlight the limitations of single testing methods and the importance of considering viral load and timing of testing when interpreting results. They also suggest the potential value of comprehensive testing approaches that combine multiple methods for more accurate diagnosis.

Consistent with previous research,^37^ 84% of individuals who had their first SARS-CoV-2 infection after vaccination developed N antibodies. In contrast to pre-Omicron studies,^17^ we observed no significant differences in N antibody detection between vaccinated and unvaccinated individuals with recent infection. Omicron’s mutations in RBD may enhance breakthrough infections by evading vaccine-induced antibody neutralization,^38–40^ potentially leading to increased reliance on and strengthening of the immune system’s N antibody response. Additionally, Omicron has a higher replication rate in the nasopharynx, saliva, and upper respiratory tract compared to ancestral variants.^41,42^ This may lead to increased shedding of viral particles, including the N protein, which the immune system can detect and generate antibodies against.

Younger individuals (<20 years) had higher S1 and RBD levels than adults over 65 years beyond 5 months post-vaccination, consistent with other studies.^43,44^ Differences associated with advancing age may be attributed to thymic involution and reduced B cell generation, differences in inflammatory response or cytokine production, and underlying comorbidities or immune modulatory medications.^45^ Among the explored comorbidities, diabetes and high triglycerides were associated with lower IgG responses. The inverse association with high triglyceride concentrations could be due to potential interference from triglycerides with the assay used. Although an interfering substances study was not performed for this study, it is a consideration for future investigations. For diabetes specifically, potential mechanisms include a reduced number of circulating helper T cells and impaired antigen presentation, as reported by Soetedjo *et al.*^46^ Further research is needed to understand the specific immunological mechanisms by which certain comorbidities compromise vaccine-induced humoral immunity. These findings emphasize the need for tailored vaccination strategies, potentially involving adjuvants to enhance immune responses in older adults and considering earlier booster doses for vulnerable populations.

This study included persons vaccinated with monovalent mRNA vaccines, initially designed against the wild-type SARS-CoV-2. While highly effective in preventing severe illness,^11,47^ these vaccines faced declining effectiveness as newer variants emerged.^48,49^ Over three-quarters of infections in our study occurred during the Omicron wave. Omicron sub-lineages from BA.1 to BA.5 and subsequent variants such as XBB.1.5 and JN.1 demonstrated increased transmissibility and immune escape due to S1 protein mutations, rendering the monovalent vaccines less protective.^50^ In response, bivalent mRNA boosters targeting both the original strain and Omicron subvariants (BA.1 or BA.4-5) were developed for use in late 2022 and demonstrated increased effectiveness against severe outcomes with Omicron infection compared to monovalent vaccines in both laboratory and real-world studies.^51–53^ COVID-19 vaccine formulations were subsequently updated for 2023–2024 to a new monovalent vaccine, designed to target the Omicron XBB.1.5 sub-lineage.^54^ The earlier bivalent and monovalent formulations are no longer authorized for use in the U.S.

Despite waning antibody levels and variant emergence, vaccines remain an important way to prevent severe illness, hospitalization, mechanical ventilation, and death associated with COVID-19.^11,53^ Additionally, effective treatments are available, further contributing to the reduction of severe outcomes. This protects individual health and reduces the economic burden on healthcare systems. Treating severe COVID-19 cases is costly, with inpatient stays averaging $11,275 as of March 2022.^55^ Widespread vaccination reduces these high-cost hospitalizations, offering clear economic benefits.

This study was subject to several limitations. First, our analysis relied on specific antibody assays measuring IgG levels of N and S1 proteins. However, other relevant immune responses, such as B cell and T cell-mediated immunity, were not assessed. Second, this study focused on antibody levels, not directly assessing their functional effectiveness against variants. Changes in levels may not always translate to altered protection against clinical outcomes. Our findings showed an association between higher anti-N IgG levels and a decreased likelihood of a subsequent positive RT-PCR test. The relationship between antibody levels and disease severity can be complex and may vary depending on factors such as the specific variant, the type of antibodies measured (IgG versus neutralizing antibodies), and individual immune response. However, our findings align with studies suggesting that higher IgG levels may be associated with a lower risk of developing symptomatic COVID-19 infection.^56,57^

Our study describes IgG responses following vaccination and infection among a cohort in Puerto Rico. Understanding the differential decay of RBD and S1 antibodies, the impact of comorbidities, and the nuances of N response following infection further informs targeted interventions and public health strategies. Some observed N and S1 serum reactivity might reflect undetected SARS-CoV-2 exposure, underscoring the value of serological testing in capturing broader population immunity compared to RT-PCR testing, which is only detectable for a short time and might miss asymptomatic or mild cases. Optimal testing to understand viral pathogenesis requires serial determinations of both viral antigens and antibody responses.

## Data Availability

Due to data security and confidentiality guidelines, all analyses, and restricted-use datasets, including questionnaire forms and code, must be requested from CDC and PMSF after submitting a concept proposal to.

## Additional Information

## Acknowledgements

COCOVID study staff and participants

## Disclaimer

The findings and conclusions in this report are those of the authors and do not necessarily represent the official position of the US Centers for Disease Control and Prevention.

## Funding

V. R.-A. acknowledges grant number U01CK000580 awarded by the Centers for Disease Control and Prevention.

## Contributions

Conception and design: ZJM, NG, VKL, LEA, CGM; Acquisition of data: CGM, DMR; Analysis and interpretation of data: ZJM, NG, VKL, DMR, JMW, LEA, GP, CGM; Drafting of the manuscript: Analysis and interpretation of data: ZJM, NG, VKL, DMR, JMW, LEA, CGM; Revision of the manuscript: All authors.

## Competing interests

The author(s) declare no competing interests.

## Notes

### Competing Interest Statement

The authors have declared no competing interest.

### Author Declarations

Approval for the COPA project was obtained from the Ponce Medical School Foundation, Inc. Institutional Review Board (protocol number 2402185168/171110-VR).

## References

1 Guevara-Hoyer, K. et al. Serological Tests in the Detection of SARS-CoV-2 Antibodies. Diagnostics (Basel) 11 (2021). 10.3390/diagnostics11040678

2 GeurtsvanKessel, C. H. et al. An evaluation of COVID-19 serological assays informs future diagnostics and exposure assessment. Nature Communications 11, 3436 (2020). 10.1038/s41467-020-17317-y

3 Ghaffari, A., Meurant, R. & Ardakani, A. COVID-19 Serological Tests: How Well Do They Actually Perform? Diagnostics (Basel) 10 (2020). 10.3390/diagnostics10070453

4 Dehgani-Mobaraki, P. et al. Long-term persistence of IgG antibodies in recovered COVID-19 individuals at 18 months post-infection and the impact of two-dose BNT162b2 (Pfizer-BioNTech) mRNA vaccination on the antibody response: Analysis using fixed-effects linear regression model. Virology 578, 111–116 (2023). 10.1016/j.virol.2022.12.003

5 Assaid, N. et al. Kinetics of SARS-CoV-2 IgM and IgG Antibodies 3 Months after COVID-19 Onset in Moroccan Patients. Am J Trop Med Hyg 108, 145–154 (2023). 10.4269/ajtmh.22-0448

6 Gilboa, M. et al. Durability of Immune Response After COVID-19 Booster Vaccination and Association With COVID-19 Omicron Infection. JAMA Netw Open 5, e2231778 (2022). 10.1001/jamanetworkopen.2022.31778

7 Yousefi, Z. et al. Long-Term Persistence of Anti-SARS-COV-2 IgG Antibodies. Curr Microbiol 79, 96 (2022). 10.1007/s00284-022-02800-0

8 Rose, R. et al. Humoral immune response after different SARS-CoV-2 vaccination regimens. BMC Medicine 20, 31 (2022). 10.1186/s12916-021-02231-x

9 Ailsworth, S. M. et al. Enhanced SARS-CoV-2 IgG durability following COVID-19 mRNA booster vaccination and comparison of BNT162b2 with mRNA-1273. Ann Allergy Asthma Immunol 130, 67–73 (2023). 10.1016/j.anai.2022.10.003

10 Adjobimey, T. et al. Comparison of IgA, IgG, and Neutralizing Antibody Responses Following Immunization With Moderna, BioNTech, AstraZeneca, Sputnik-V, Johnson and Johnson, and Sinopharm’s COVID-19 Vaccines. Front Immunol 13, 917905 (2022). 10.3389/fimmu.2022.917905

11 Song, S., Madewell, Z. J., Liu, M., Longini, I. M. & Yang, Y. Effectiveness of SARS-CoV-2 vaccines against Omicron infection and severe events: a systematic review and meta-analysis of test-negative design studies. Front Public Health 11, 1195908 (2023). 10.3389/fpubh.2023.1195908

12 Virk, A. et al. Hybrid Immunity Provides Protective Advantage Over Vaccination or Prior Remote Coronavirus Disease 2019 Alone. Open Forum Infect Dis 10, ofad161 (2023). 10.1093/ofid/ofad161

13 McBride, R., van Zyl, M. & Fielding, B. C. The coronavirus nucleocapsid is a multifunctional protein. Viruses 6, 2991–3018 (2014). 10.3390/v6082991

14 Huang, Y., Yang, C., Xu, X. F., Xu, W. & Liu, S. W. Structural and functional properties of SARS-CoV-2 spike protein: potential antivirus drug development for COVID-19. Acta Pharmacol Sin 41, 1141–1149 (2020). 10.1038/s41401-020-0485-4

15 Tai, W. et al. Characterization of the receptor-binding domain (RBD) of 2019 novel coronavirus: implication for development of RBD protein as a viral attachment inhibitor and vaccine. Cell Mol Immunol 17, 613–620 (2020). 10.1038/s41423-020-0400-4

16 Pang, N. Y., Pang, A. S., Chow, V. T. & Wang, D. Y. Understanding neutralising antibodies against SARS-CoV-2 and their implications in clinical practice. Mil Med Res 8, 47 (2021). 10.1186/s40779-021-00342-3

17 Follmann, D. et al. Antinucleocapsid Antibodies After SARS-CoV-2 Infection in the Blinded Phase of the Randomized, Placebo-Controlled mRNA-1273 COVID-19 Vaccine Efficacy Clinical Trial. Ann Intern Med 175, 1258–1265 (2022). 10.7326/m22-1300

18 Pooley, N. et al. Durability of Vaccine-Induced and Natural Immunity Against COVID-19: A Narrative Review. Infect Dis Ther 12, 367–387 (2023). 10.1007/s40121-022-00753-2

19 Townsend, J. P., Hassler, H. B., Sah, P., Galvani, A. P. & Dornburg, A. The durability of natural infection and vaccine-induced immunity against future infection by SARS-CoV-2. Proc Natl Acad Sci U S A 119, e2204336119 (2022). 10.1073/pnas.2204336119

20 Hornsby, H. et al. Omicron infection following vaccination enhances a broad spectrum of immune responses dependent on infection history. Nat Commun 14, 5065 (2023). 10.1038/s41467-023-40592-4

21 Canetti, M. et al. Immunogenicity and efficacy of fourth BNT162b2 and mRNA1273 COVID-19 vaccine doses; three months follow-up. Nature Communications 13, 7711 (2022). 10.1038/s41467-022-35480-2

22 Adams, L. E. et al. Risk factors for infection with chikungunya and Zika viruses in southern Puerto Rico: A community-based cross-sectional seroprevalence survey. PLoS Negl Trop Dis 16, e0010416 (2022). 10.1371/journal.pntd.0010416

23 Rodríguez, D. M. et al. HTrack: A new tool to facilitate public health field visits and electronic data capture. PLoS One 15, e0244028 (2020). 10.1371/journal.pone.0244028

24 Major, C. G., et al. Investigating SARS-CoV-2 Incidence and Morbidity in Ponce, Puerto Rico: Protocol and Baseline Results From a Community Cohort Study. JMIR Res Protoc 13, e53837 (2024). 10.2196/53837

25 Madewell, Z. J. et al. Diagnostic Accuracy of the Abbott BinaxNOW COVID-19 Antigen Card Test, Puerto Rico. Influenza Other Respir Viruses 18, e13305 (2024). 10.1111/irv.13305

26 Luminex. xMAP® SARS-CoV-2 Antibody Testing, <https://www.luminexcorp.com/xmap-sars-cov-2-antibody-testing/> (2023).

27 Luminex. xMAP® SARS-CoV-2 Multi-Antigen IgG Assay Package Insert, <https://www.fda.gov/media/140256/download> (2022).

28 Centers for Disease Control and Prevention. Investigative Criteria for Suspected Cases of SARS-CoV-2 Reinfection (ICR), <https://stacks.cdc.gov/view/cdc/96072> (2020).

29 Santiago, G. A. et al. SARS-CoV-2 Omicron Replacement of Delta as Predominant Variant, Puerto Rico. Emerging Infectious Diseases 29, 855 (2023).

30 Santiago, G. A. et al. Genomic surveillance of SARS-CoV-2 in Puerto Rico enabled early detection and tracking of variants. Commun Med (Lond) 2, 100 (2022). 10.1038/s43856-022-00168-7

31 Jones, J. M. et al. Estimates of SARS-CoV-2 Seroprevalence and Incidence of Primary SARS-CoV-2 Infections Among Blood Donors, by COVID-19 Vaccination Status - United States, April 2021-September 2022. MMWR Morb Mortal Wkly Rep 72, 601–605 (2023). 10.15585/mmwr.mm7222a3

32 Lapuente, D., Winkler, T. H. & Tenbusch, M. B-cell and antibody responses to SARS-CoV-2: infection, vaccination, and hybrid immunity. Cell Mol Immunol 21, 144–158 (2024). 10.1038/s41423-023-01095-w

33 Wagner, A. et al. Neutralising SARS-CoV-2 RBD-specific antibodies persist for at least six months independently of symptoms in adults. Communications Medicine 1, 13 (2021). 10.1038/s43856-021-00012-4

34 Upadhyay, V., Lucas, A., Panja, S., Miyauchi, R. & Mallela, K. M. G. Receptor binding, immune escape, and protein stability direct the natural selection of SARS-CoV-2 variants. J Biol Chem 297, 101208 (2021). 10.1016/j.jbc.2021.101208

35 Hellewell, J. et al. Estimating the effectiveness of routine asymptomatic PCR testing at different frequencies for the detection of SARS-CoV-2 infections. BMC Med 19, 106 (2021). 10.1186/s12916-021-01982-x

36 Ko, J. H. et al. Neutralizing Antibody Production in Asymptomatic and Mild COVID-19 Patients, in Comparison with Pneumonic COVID-19 Patients. J Clin Med 9 (2020). 10.3390/jcm9072268

37 den Hartog, G., et al. Assessment of hybrid population immunity to SARS-CoV-2 following breakthrough infections of distinct SARS-CoV-2 variants by the detection of antibodies to nucleoprotein. Scientific Reports 13, 18394 (2023). 10.1038/s41598-023-45718-8

38 Cao, Y. et al. Omicron escapes the majority of existing SARS-CoV-2 neutralizing antibodies. Nature 602, 657–663 (2022). 10.1038/s41586-021-04385-3

39 Wang, Q. et al. Antibody evasion by SARS-CoV-2 Omicron subvariants BA.2.12.1, BA.4 and BA.5. Nature 608, 603–608 (2022). 10.1038/s41586-022-05053-w

40 Planas, D. et al. Resistance of Omicron subvariants BA.2.75.2, BA.4.6, and BQ.1.1 to neutralizing antibodies. Nat Commun 14, 824 (2023). 10.1038/s41467-023-36561-6

41 Granerud, B. K. et al. Omicron Variant Generates a Higher and More Sustained Viral Load in Nasopharynx and Saliva Than the Delta Variant of SARS-CoV-2. Viruses 14 (2022). 10.3390/v14112420

42 Hui, K. P. Y. et al. Replication of SARS-CoV-2 Omicron BA.2 variant in ex vivo cultures of the human upper and lower respiratory tract. EBioMedicine 83, 104232 (2022). 10.1016/j.ebiom.2022.104232

43 Karachaliou, M. et al. SARS-CoV-2 infection, vaccination, and antibody response trajectories in adults: a cohort study in Catalonia. BMC Medicine 20, 347 (2022). 10.1186/s12916-022-02547-2

44 Richards, N. E. et al. Comparison of SARS-CoV-2 Antibody Response by Age Among Recipients of the BNT162b2 vs the mRNA-1273 Vaccine. JAMA Netw Open 4, e2124331 (2021). 10.1001/jamanetworkopen.2021.24331

45 Bartleson, J. M. et al. SARS-CoV-2, COVID-19 and the aging immune system. Nature Aging 1, 769–782 (2021). 10.1038/s43587-021-00114-7

46 Soetedjo, N. N. M., Iryaningrum, M. R., Lawrensia, S. & Permana, H. Antibody response following SARS-CoV-2 vaccination among patients with type 2 diabetes mellitus: A systematic review. Diabetes Metab Syndr 16, 102406 (2022). 10.1016/j.dsx.2022.102406

47 Ssentongo, P. et al. SARS-CoV-2 vaccine effectiveness against infection, symptomatic and severe COVID-19: a systematic review and meta-analysis. BMC Infect Dis 22, 439 (2022). 10.1186/s12879-022-07418-y

48 Madewell, Z. J., Yang, Y., Longini, I. M., Jr., Halloran, M. E. & Dean, N. E. Household Secondary Attack Rates of SARS-CoV-2 by Variant and Vaccination Status: An Updated Systematic Review and Meta-analysis. JAMA Netw Open 5, e229317 (2022). 10.1001/jamanetworkopen.2022.9317

49 Menegale, F. et al. Evaluation of Waning of SARS-CoV-2 Vaccine-Induced Immunity: A Systematic Review and Meta-analysis. JAMA Netw Open 6, e2310650 (2023). 10.1001/jamanetworkopen.2023.10650

50 Chatterjee, S., Bhattacharya, M., Nag, S., Dhama, K. & Chakraborty, C. A Detailed Overview of SARS-CoV-2 Omicron: Its Sub-Variants, Mutations and Pathophysiology, Clinical Characteristics, Immunological Landscape, Immune Escape, and Therapies. Viruses 15 (2023). 10.3390/v15010167

51 Hyun, H. J. et al. Neutralizing Activity against BQ.1.1, BN.1, and XBB.1 in Bivalent COVID-19 Vaccine Recipients: Comparison by the Types of Prior Infection and Vaccine Formulations. Vaccines (Basel) 11 (2023). 10.3390/vaccines11081320

52 Davis-Gardner, M. E. et al. Neutralization against BA.2.75.2, BQ.1.1, and XBB from mRNA Bivalent Booster. N Engl J Med 388, 183–185 (2023). 10.1056/NEJMc2214293

53 Song, S. et al. A systematic review and meta-analysis on the effectiveness of bivalent mRNA booster vaccines against Omicron variants. Vaccine (2024). 10.1016/j.vaccine.2024.04.049

54 Regan, J. J. et al. Use of Updated COVID-19 Vaccines 2023-2024 Formula for Persons Aged ≥6 Months: Recommendations of the Advisory Committee on Immunization Practices - United States, September 2023. MMWR Morb Mortal Wkly Rep 72, 1140–1146 (2023). 10.15585/mmwr.mm7242e1

55 Kapinos, K. A. et al. Inpatient Costs of Treating Patients With COVID-19. JAMA Netw Open 7, e2350145 (2024). 10.1001/jamanetworkopen.2023.50145

56 Messchendorp, A. L. et al. Incidence and Severity of COVID-19 in Relation to Anti-Receptor-Binding Domain IgG Antibody Level after COVID-19 Vaccination in Kidney Transplant Recipients. Viruses 16 (2024). 10.3390/v16010114

57 Chen, X. et al. Association between levels of IgG antibodies from vaccines and Omicron symptomatic infection among children and adolescents in China. Front Med (Lausanne) 10, 1240340 (2023). 10.3389/fmed.2023.1240340

