## Supplement for "Durability of SARS-CoV-2 IgG Antibodies: Insights from a Longitudinal Study, Puerto Rico"

**Figure S1.** Seropositivity of SARS-CoV-2 IgG S1 antibody responses (top) and log(MFI) for S1 (bottom) by days since vaccination among participants who did not have a previous positive RT-PCR and were N seronegative, Puerto Rico, 2020–2022.

**Figure S2.** Seropositivity of SARS-CoV-2 IgG RBD antibody responses (top) and log(MFI) for RBD (bottom) by days since vaccination among participants who did not have a previous positive RT-PCR and were N seronegative, Puerto Rico, 2020–2022.

**Figure S3.** SARS-CoV-2 IgG antibody responses by vaccination status, days since last vaccine, and previous RT-PCR positivity, Puerto Rico, 2020–2022.

**Figure S4.** Correlations between SARS-CoV-2 IgG antibody responses by vaccination status, Puerto Rico, 2020–2022. Pearson’s correlation coefficients (95% CIs) are shown.

**Figure S5.** SARS-CoV-2 IgG antibody responses by vaccination status, days since last vaccine, and age group among participants who did not have a previous positive RT-PCR and were N seronegative, Puerto Rico, 2020–2022.

**Figure S6.** Unadjusted and adjusted associations between time since last vaccine and time since RT-PCR positivity, and SARS-CoV-2 IgG antibody responses, Puerto Rico, 2020–2022.

**Figure S7.** Paired SARS-CoV-2 IgG antibody responses at two time points six months apart following a second dose for individuals without a previous positive RT-PCR test, Puerto Rico, 2020–2022.

**Figure S8.** SARS-CoV-2 IgG antibody responses by sequence of vaccination (V) and positive RT-PCR confirmed infection (I) among individuals who received two doses, Puerto Rico, 2020–2022.

**Figure S9.** Unadjusted and adjusted associations between chronic conditions and SARS-CoV-2 IgG antibody responses, Puerto Rico, 2020–2022.

**Table S1.** Participation per milestone visit. Decrease in milestone attendance attributed to general loss to follow-up, participation in weekly PCR-test falling below 80% attendance, or lack of serology data, Puerto Rico, 2020–2022.

**Table S2.** Vaccination status recorded on final visit for COCOVID participants by vaccine manufacturer for those who had received at least one dose, Puerto Rico, 2020–2022.

**Appendix S1.** Supplemental Methods.

**Figure S1.** Seropositivity of SARS-CoV-2 IgG S1 antibody responses (top) and log(MFI) for S1 (bottom) by days since vaccination among participants who did not have a previous positive RT-PCR and were N seronegative, Puerto Rico, 2020–2022. Each color represents a specific time point post-vaccination.

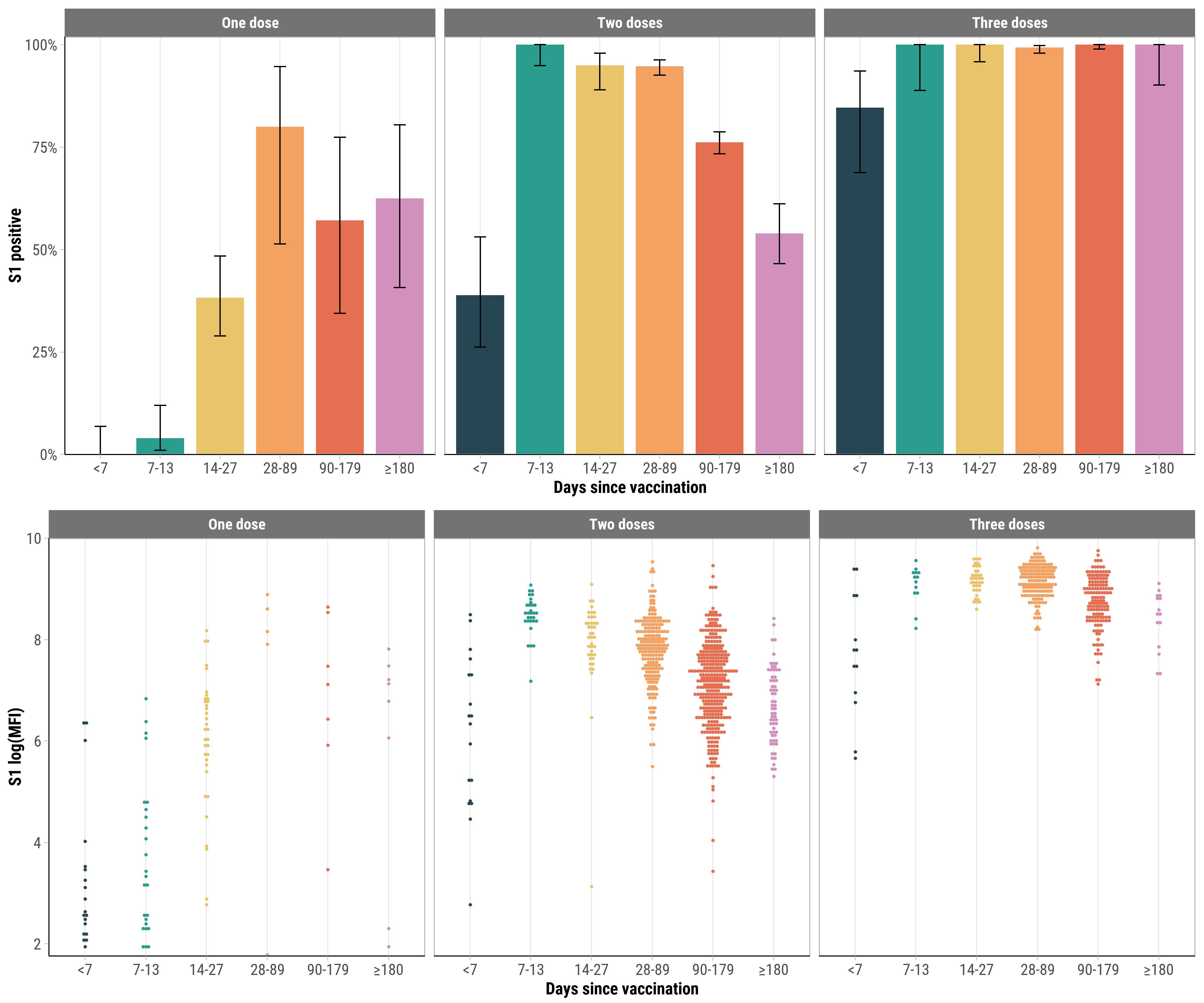

**Figure S2.** Seropositivity of SARS-CoV-2 IgG RBD antibody responses (top) and log(MFI) for RBD (bottom) by days since vaccination among participants who did not have a previous positive RT-PCR and were N seronegative, Puerto Rico, 2020–2022. Each color represents a specific time point post-vaccination.

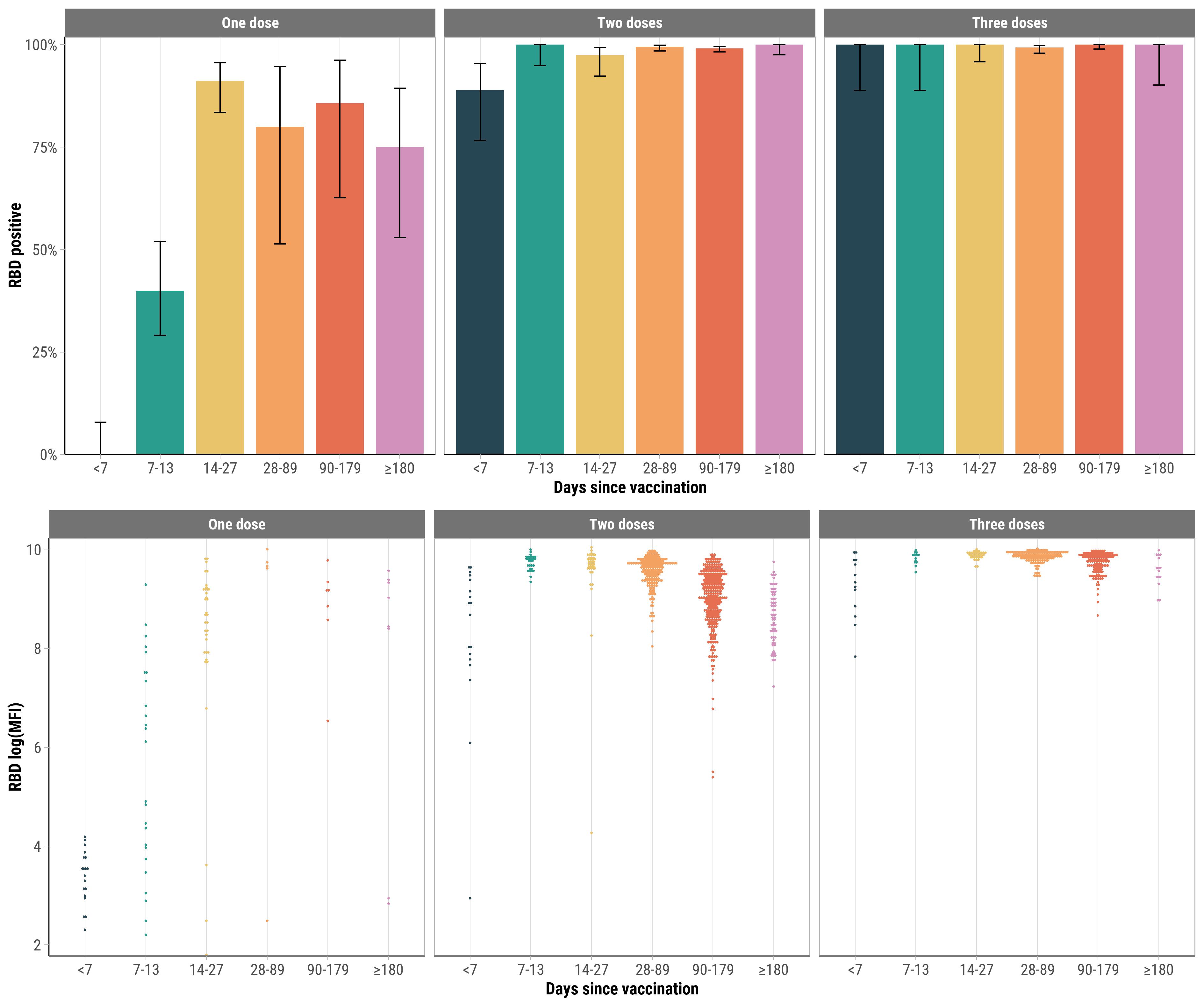

**Figure S3.** SARS-CoV-2 IgG antibody responses by vaccination status, days since last vaccine, and previous RT-PCR positivity, Puerto Rico, 2020–2022.

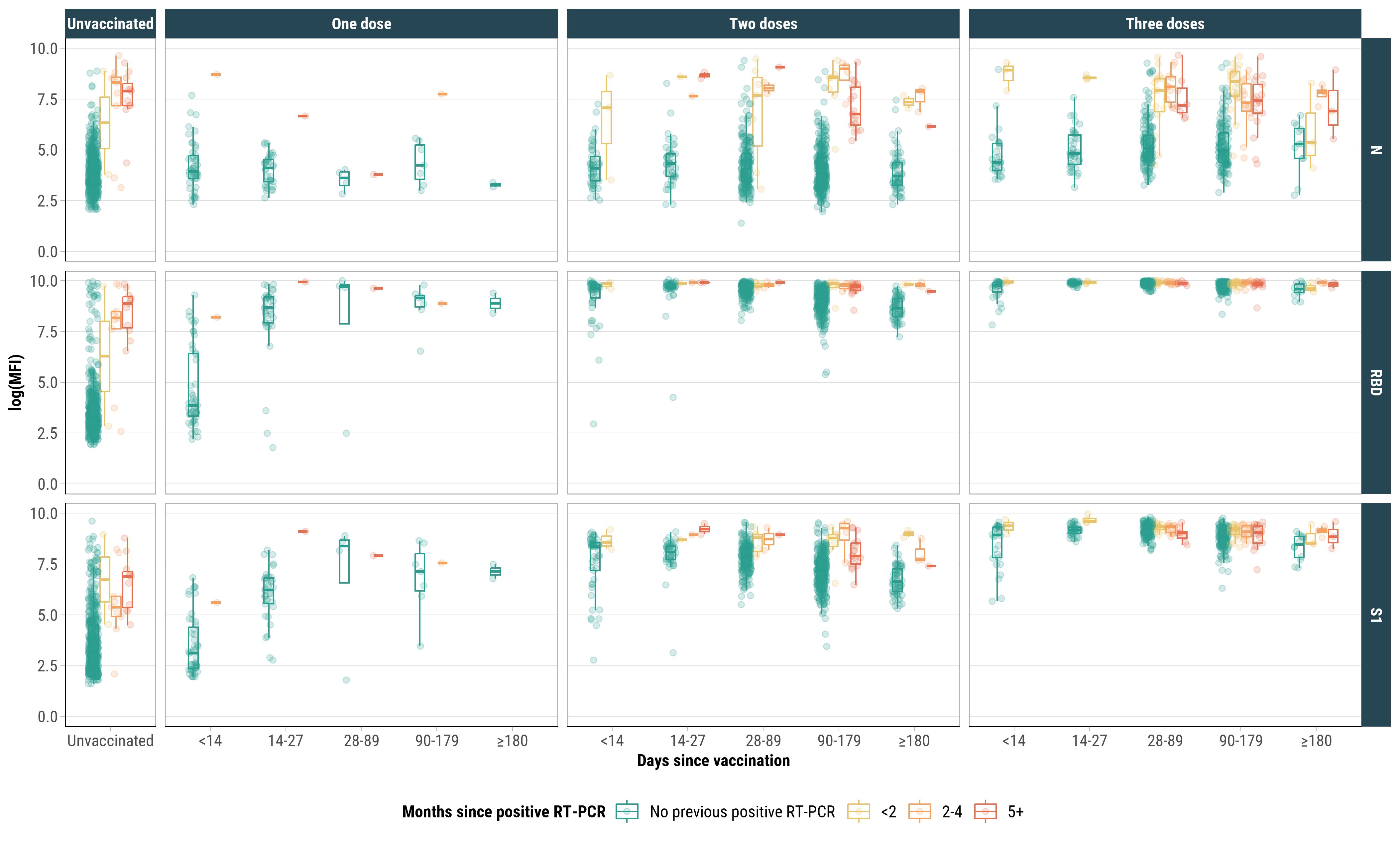

**Figure S4.** Correlations between SARS-CoV-2 IgG antibody responses by vaccination status, Puerto Rico, 2020–2022. Pearson’s correlation coefficients (95% CIs) are shown.

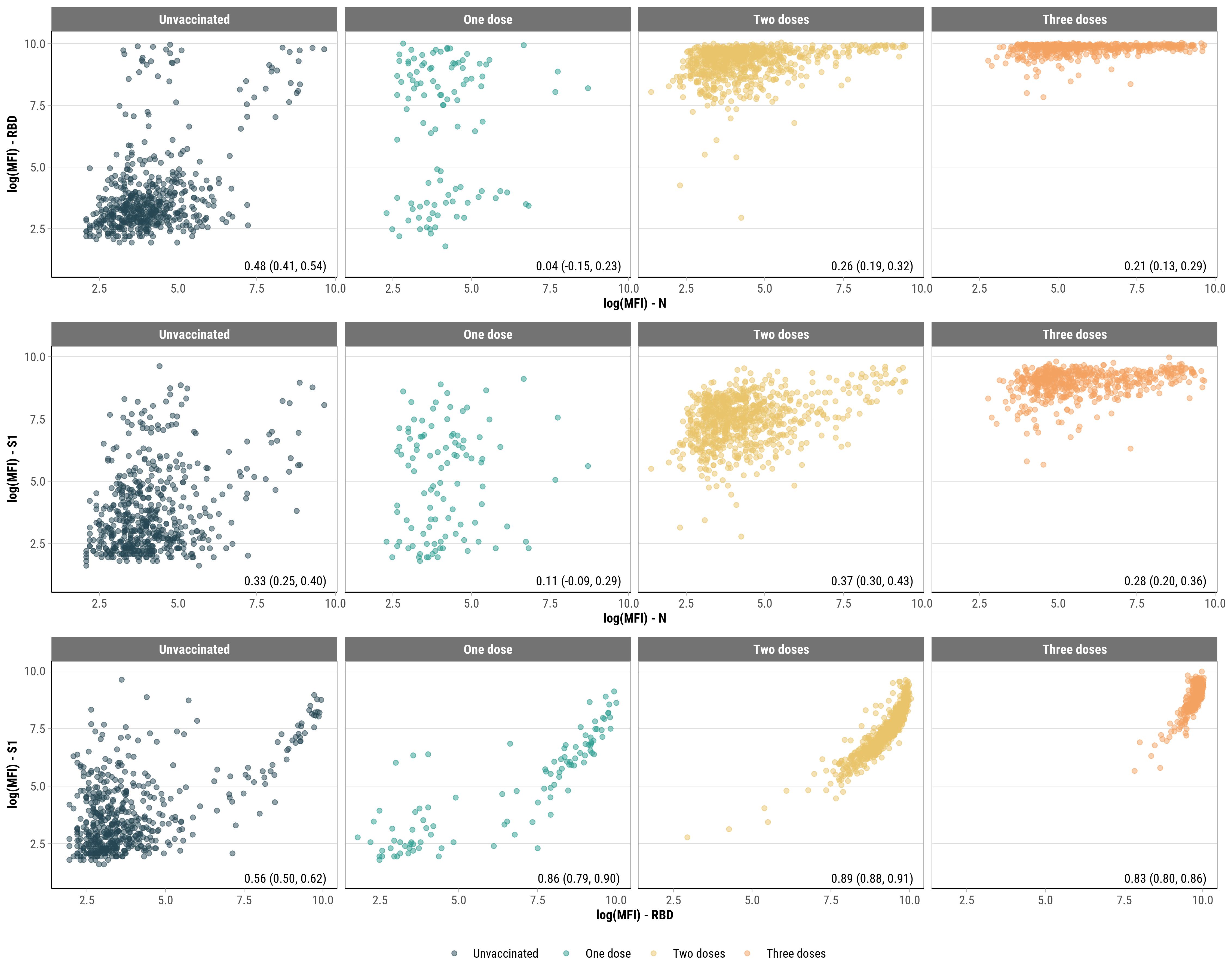

**Figure S5**. SARS-CoV-2 IgG antibody responses by vaccination status, days since last vaccine, and age group among participants who did not have a previous positive RT-PCR and were N seronegative, Puerto Rico, 2020–2022.

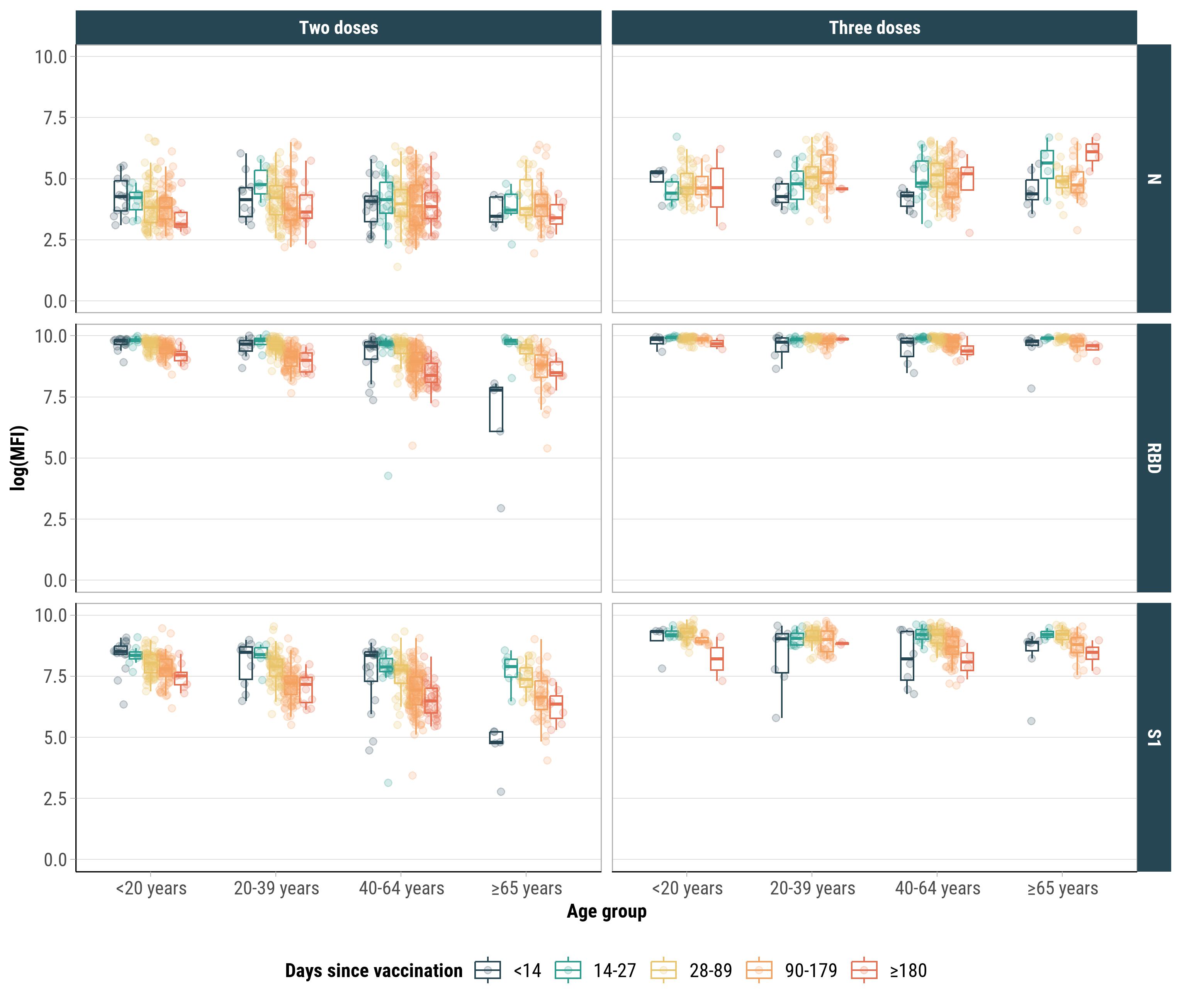

**Figure S6.** Unadjusted and adjusted associations between time since last vaccine and time since RT-PCR positivity, and SARS-CoV-2 IgG antibody responses, Puerto Rico, 2020–2022.

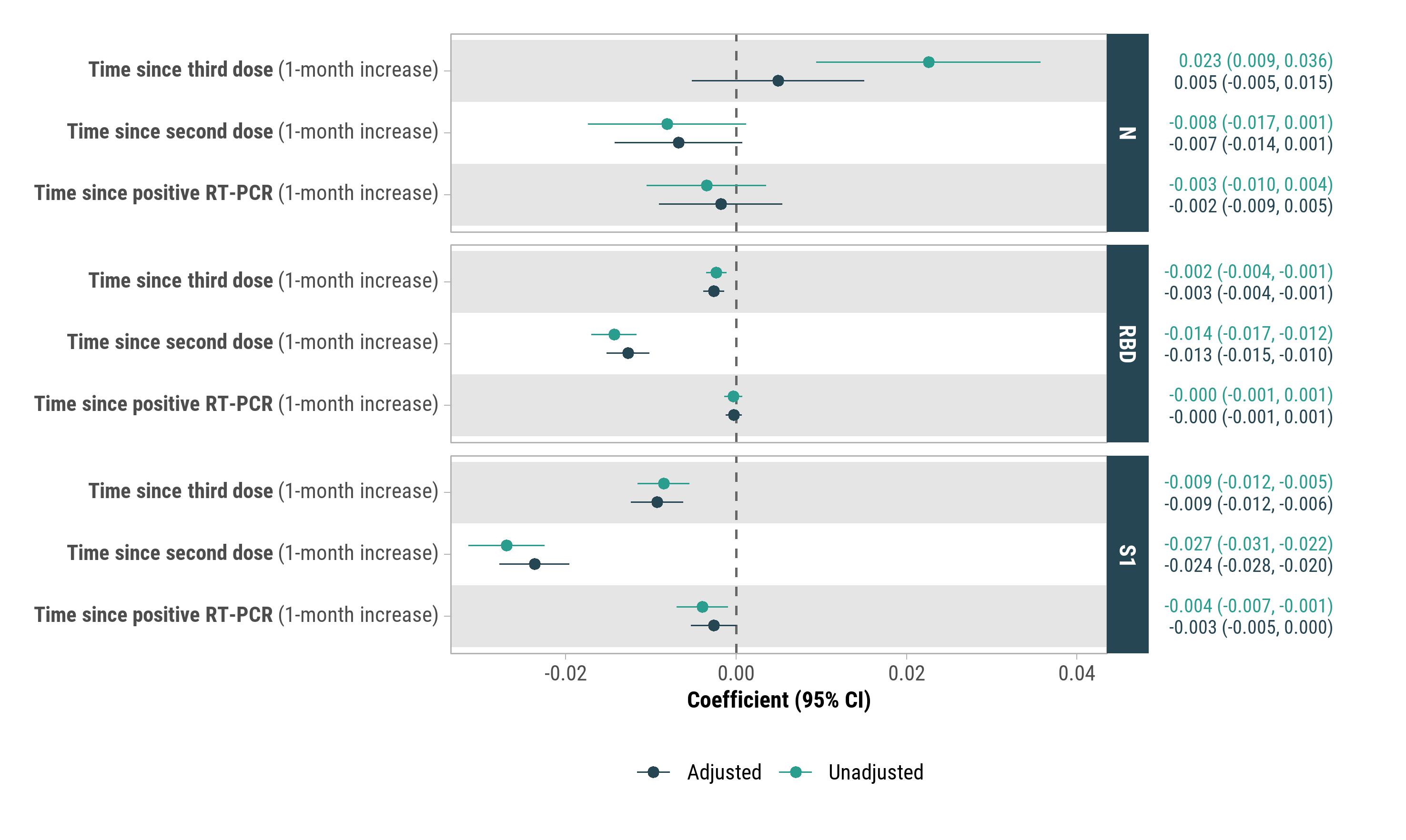

Associations with each antibody (N, RBD, S1) were evaluated using separate generalized linear mixed-effects models with a log link function, treating continuous antibody responses as the outcome variable. Multivariate models were adjusted for age group, sex, and any chronic condition. Associations with time since infection were also adjusted for vaccination status, while associations with time since vaccination were adjusted for RT-PCR positivity. β coefficients and 95% confidence intervals are shown.

**Figure S7.** Paired SARS-CoV-2 IgG antibody responses at two time points six months apart following a second dose for individuals without a previous positive RT-PCR test, Puerto Rico, 2020–2022.

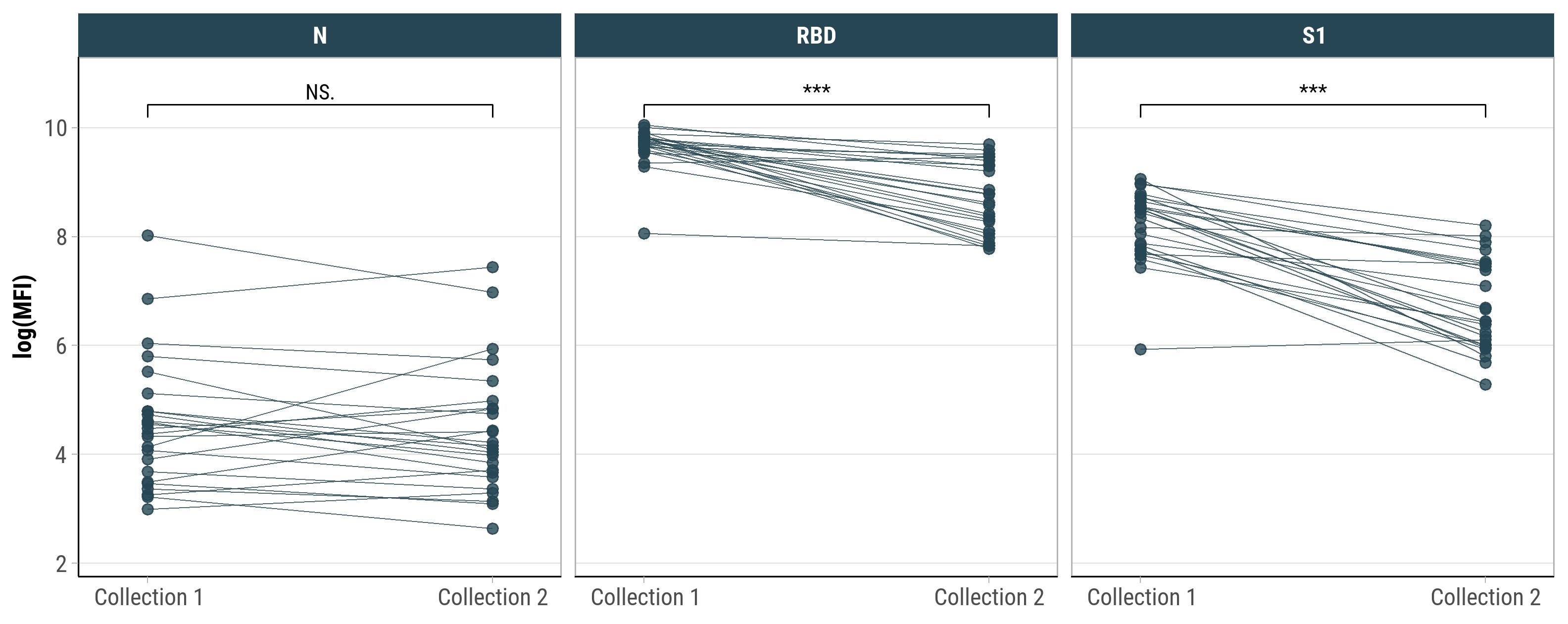

****P* < 0.001, NS: not significant. There were no participants with paired SARS-CoV-2 IgG antibody responses at two time points six months apart with a second dose and a previous positive RT-PCR.

**Figure S8.** SARS-CoV-2 IgG antibody responses by sequence of vaccination (V) and positive RT-PCR confirmed infection (I) among individuals who received two doses, Puerto Rico, 2020–2022.

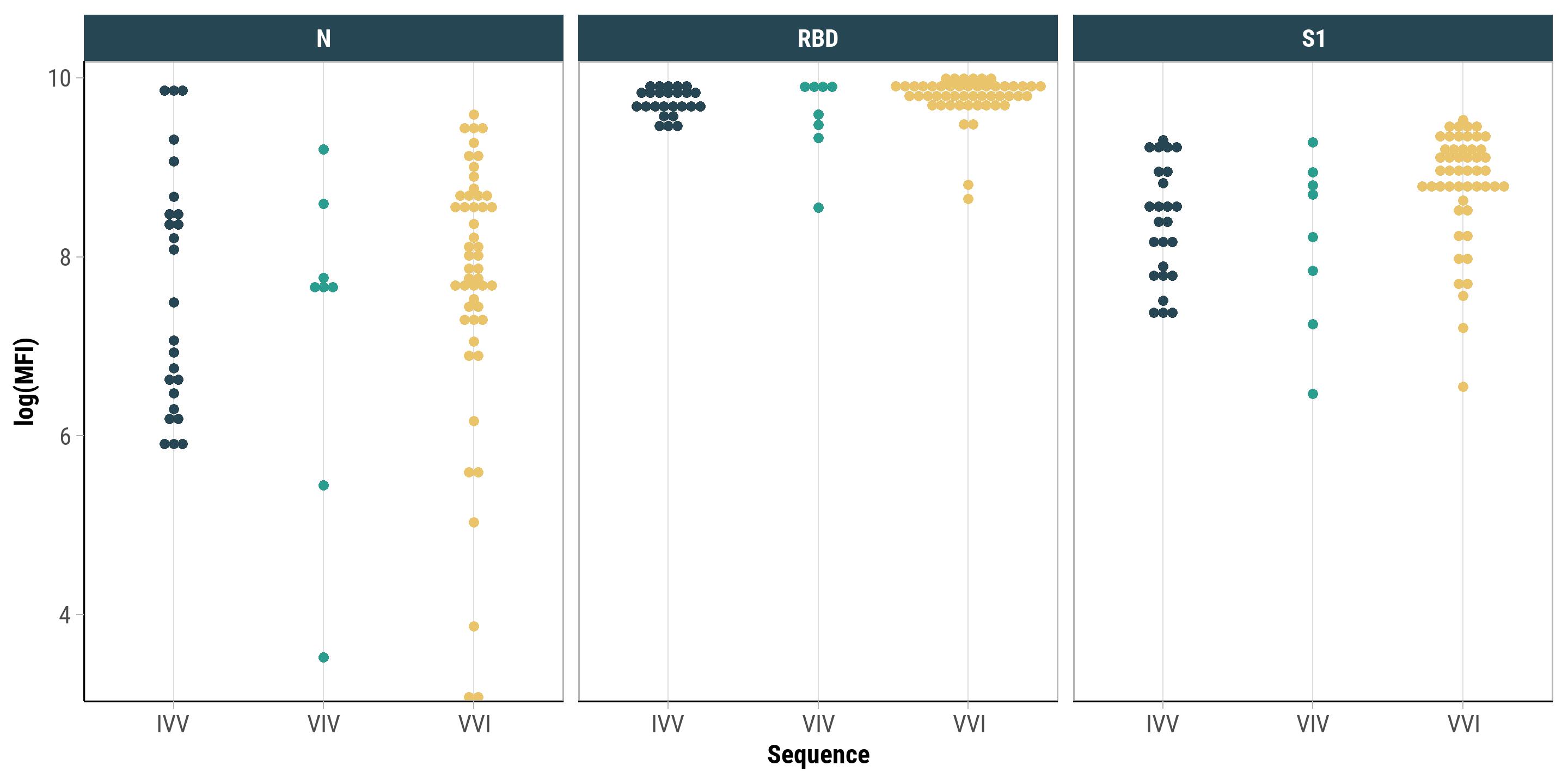

E.g., IVV includes participants infected, vaccinated, and vaccinated, in that order. For IVV, median days since last positive RT-PCR were 55 (IQR: 30, 69). For VVI, median days since last vaccine was 134 (IQR: 93, 150). For VIV, median days since last vaccine was 28 (IQR: 24, 99). **Figure S9.** Unadjusted and adjusted associations between chronic conditions and SARS-CoV-2 IgG antibody responses, Puerto Rico, 2020–2022.

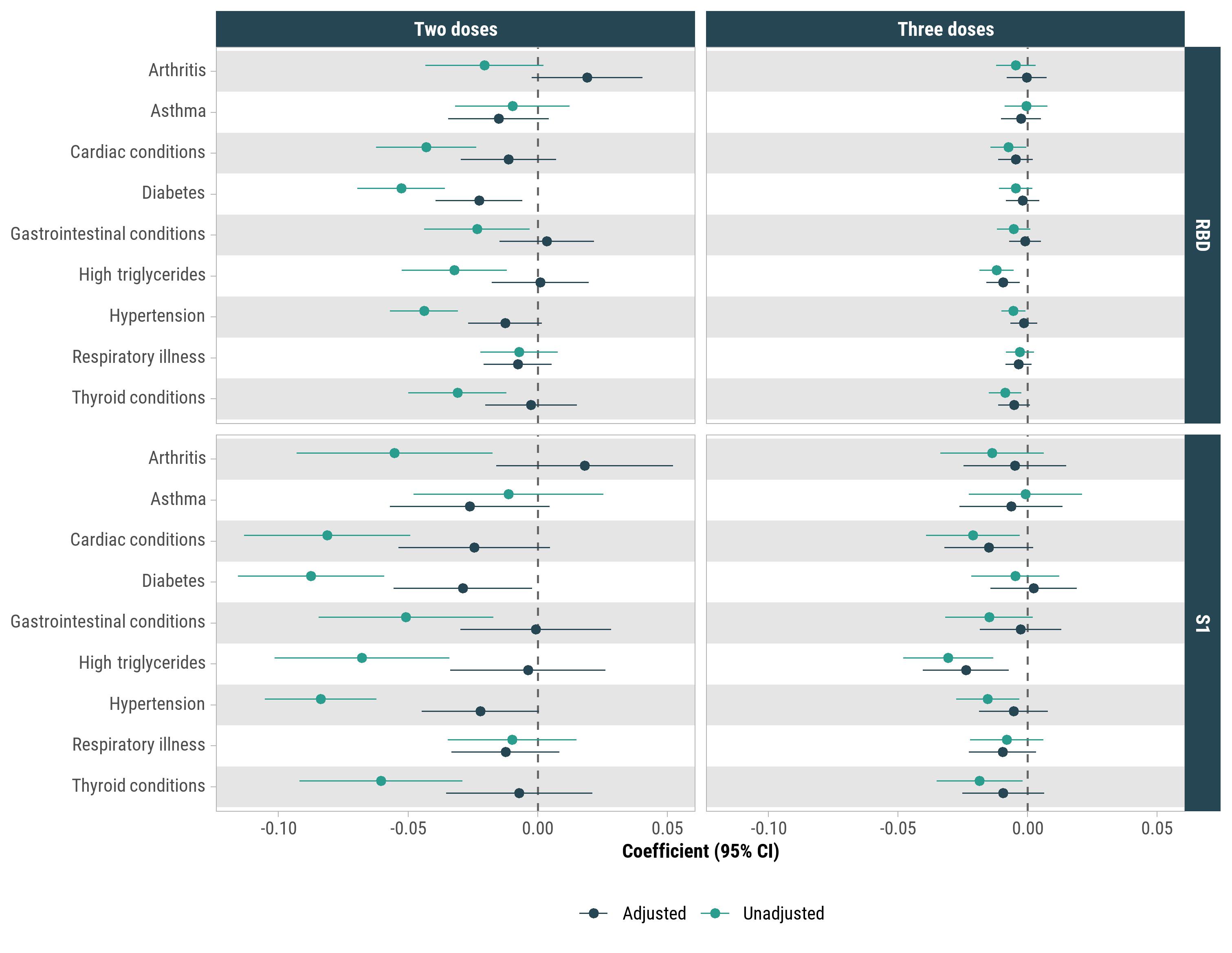

Associations between each chronic condition and each antibody (RBD, S1) were evaluated using separate generalized linear mixed-effects models with a log link function, treating continuous antibody responses as the outcome variable. Multivariable models were adjusted for RT-PCR positivity, age group, sex, and time since last vaccine. β coefficients and 95% confidence intervals are shown.

| **Table S1. Participation per milestone visit.** Decrease in milestone attendance attributed to general loss to follow-up, participation in weekly PCR-test falling below 80% attendance, or lack of serology data, Puerto Rico, 2020–2022. | | | | | | | |
| --- | --- | --- | --- | --- | --- | --- | --- |
| **Milestone** | **Observations** | **Consistent PCR Testing** | **Inconsistent PCR Testing AND Positive PCR Test** | **Invalid**  **Serum** | **Invalid**  **Vaccine** | **Inconsistent PCR Testing AND Non-positive PCR Test** | **Loss to**  **Follow-up** |
| **Baseline** | 1027 | - | - | - | - | - | - |
| **6 months** | 789 | 789 | 0 | 99 | 8 | 24 | 107 |
| **12 months** | 774 | 772 | 2 | 32 | 10 | 55 | 156 |
| **18 months** | 567 | 505 | 62 | 33 | 4 | 174 | 249 |

| **Table S2**. Vaccination status recorded on final visit for COCOVID participants by vaccine manufacturer for those who had received at least one dose, Puerto Rico, 2020–2022. | |
| --- | --- |
| Vaccination status | N = 796  n (%) |
| **One dose** (N = 10) |  |
| BNT162b2 | 9 (1.1) |
| mRNA-1273 | 1 (0.1) |
| **Two doses** (N = 316) |  |
| BNT162b2 / BNT162b2 | 216 (27.1) |
| mRNA-1273 / mRNA-1273 | 97 (12.2) |
| BNT162b2 / mRNA-1273 | 2 (0.3) |
| **Three doses** (N = 468) |  |
| BNT162b2 / BNT162b2 / BNT162b2 | 237 (29.8) |
| mRNA-1273 / mRNA-1273 / mRNA-1273 | 144 (18.1) |
| BNT162b2 / BNT162b2 / unknown | 71 (8.9) |
| mRNA-1273 / mRNA-1273 / BNT162b2 | 5 (0.6) |
| mRNA-1273 / mRNA-1273 / unknown | 4 (0.5) |
| mRNA-1273 / BNT162b2 / unknown | 4 (0.5) |
| BNT162b2 / BNT162b2 / mRNA-1273 | 3 (0.4) |
| **Four doses** (N = 2) |  |
| mRNA-1273 / mRNA-1273 / mRNA-1273 / mRNA-1273 | 2 (0.3) |

**Appendix S1**

*Supplemental Methods*

Generalized linear mixed-effects regression was used to evaluate unadjusted and adjusted associations between time since infection (one-month intervals), time since vaccination (one-month intervals), and chronic conditions (arthritis, asthma, cardiac conditions, diabetes, gastrointestinal conditions, high triglycerides, hypertension, respiratory illness, thyroid conditions), and SARS-CoV-2 IgG antibody responses (N, RBD, S1). Generalized linear models with a log link function were used to model continuous antibody responses, accounting for overdispersion and potential non-linear relationships between predictors and antibody levels. Associations with each antibody (N, RBD, S1) were evaluated in separate regression models. Multivariate models were adjusted for age group, sex, and any chronic condition as fixed effects, and participant ID as a random intercept. Associations with time since infection were also adjusted for vaccination status. Associations with time since vaccination were also adjusted for RT-positivity. Separate models, adjusted for age group, sex, previous RT-PCR positivity, and time since last vaccine were used to examine the associations between chronic conditions and IgG responses in individuals who completed two and three doses.

Mixed-effects logistic regression was used to analyze unadjusted and adjusted associations between antibody seroreactivity and RT-PCR positivity in the subsequent six months. Seroreactivity is a binary outcome (positive or negative), thus logistic regression was used. For this analysis, antibody tests were included if they occurred during the more contemporary Omicron predominant period when vaccination was more widespread. RBD was excluded given that almost all participants were RBD seropositive during this period. Multivariate models were adjusted for age group, sex, and any chronic condition as fixed effects, and participant ID as a random intercept. In addition to evaluating binary N and S1 seroreactivity, we also evaluated associations between subsequent RT-PCR positivity and log(MFI) for N and S1 categorized as ≥8.56, 7.56-8.55, 6.56-7.55, and seronegative. The seropositivity threshold for MFI is approximately ln(700) = 6.54, and these categories were chosen with increments of 1 above this threshold to capture potential trends in the associations with subsequent RT-PCR positivity. Generalized variance inflation factors were used to evaluate collinearity between independent variables in the adjusted models. Values of P<0.05 were considered statistically significant.
